# The interaction between influenza vaccination and nasal pneumococcal colonization alters airway T cell responses and alveolar macrophage activation

**DOI:** 10.64898/2026.02.05.26345662

**Authors:** Yuzhou Evelyn Tong, Sergio Triana, Daniela D. Russo, Jesus Reine, Jamie Rylance, Simon P. Jochems, Oluwasefunmi Akeju, Pardis C. Sabeti, Alex K. Shalek, Daniela M. Ferreira, Elena Mitsi

## Abstract

**Background:** Influenza vaccination and bacterial colonization both shape immunity in the respiratory tract, yet their combined impact on the human lung mucosa remains poorly understood. Secondary bacterial pneumonia following influenza infection is a leading cause of mortality, underscoring the need to define how vaccines and microbes intersect at the airway interface.

**Methods:** Using the Experimental Human Pneumococcal Challenge (EHPC) model, we examined how intramuscular inactivated (TIV) and nasal live attenuated (LAIV) influenza vaccines, with or without *Streptococcus pneumoniae* colonization, modulate lower airway immunity. Bronchoalveolar lavage samples from 22 adults were profiled by single-cell RNA-seq (>40,000 cells), flow cytometry, cytokine multiplexing, and macrophage functional assays.

**Findings:** LAIV recipients who became colonized with *S. pneumoniae* displayed heightened influenza-specific CD4⁺ T cell responses and enhanced alveolar macrophage (AM) opsonophagocytic activity, showing that nasal bacterial colonization can act as natural mucosal adjuvant. Single-cell transcriptomics revealed four AM gene modules; among them, an interferon-driven “anti-microbial” program correlated with enhanced phagocytosis, whereas a complement- and antigen-presentation module associated with IFNγ-iNOS/ROS signaling was attenuated in colonized vaccinees. Given that AMs are poor antigen-presenting cells, this shift likely reflects reprogramming toward cytokine-mediated immune modulation rather than direct T cell activation. The elevated influenza-specific CD4⁺ T cell responses may instead represent feedback from enhanced local activation. Together, these data indicate that vaccination and colonization synergize to rewire AM-T cell communication, fine-tuning anti-viral and anti-bacterial defenses. Similar transcriptional perturbations in public COVID-19 and lung cancer datasets underscore the broader relevance of these macrophage modules across lung disease contexts.

**Conclusions:** Our findings define how influenza vaccination and pneumococcal colonization converge in the human lung to reprogram AM-T cell crosstalk, enhancing local immune responses and protective immunity. By uncovering conserved macrophage modules and mechanisms that shape mucosal defense, this study provides a framework for designing next-generation respiratory vaccines and strategies to mitigate post-viral bacterial pneumonia.

## Introduction

Pneumonia remains a significant global health concern, with post-influenza secondary pneumococcal pneumonia emerging as a leading cause of mortality worldwide^1^. Colonization of the human nasopharynx with *Streptococcus pneumoniae* (Spn, pneumococcus), known as pneumococcal carriage, is considered a prerequisite for developing pneumonia. However, it is a common event (20-90% prevalence in children <5 years), and also an essential step in developing pneumococcal immunity, both systemically and across the respiratory mucosa^2,3^. Pneumococcal disease is most prevalent in children below the age of 5 and the elderly^4,5^. In both age groups, co-infection with influenza virus often worsens the disease outcome, leading to increased hospitalisation rates, mortality and morbidity. Currently, there exists two main influenza vaccination strategies, each inducing distinct immunological profiles. Live attenuated influenza vaccine (LAIV, an intranasally administered cold-adapted vaccine that replicates transiently in the nose), tend to be used more frequently in children due to its pain-free delivery and the potential to elicit both humoral and cellular immune responses, mimicking natural infection induced immunity in the absence of disease. During co-infection with pneumococcus, studies have shown that it can delay clearance of secondary bacterial colonization by compromising neutrophil functions and impairing monocyte recruitment in the nasal mucosa^6,7^. On the other hand, Intramuscularly delivered Tetravalent Influenza Vaccines (TIVs, a type of Inactivated Influenza vaccine) are most common in the US across adult and pediatric populations, and more effectively generate serum antibodies, offering a “boost” to an immune system with already extensive pre-existing influenza immunity^8^. These distinct vaccine platforms offer a unique opportunity to investigate how mucosal versus systemic immune priming by influenza vaccination alters susceptibility to pneumococcal infection and shapes the immunological landscape of the distal lung.

The progression from asymptomatic nasopharyngeal colonization to pneumococcal pneumonia involves a complex process of bacterial translocation that is not fully understood. During pneumococcal colonization, both in the absence and presence of disease, pneumococcal aspiration drives movement to the lower respiratory tract. This is supported by recent human studies which demonstrate that bacteria present in the lung of pneumonia patients originate from the nasopharynx, with whole genome sequencing showing >99% sequence similarity between isolates from both sites in the same patient^9^. The aspiration process is facilitated by increased pneumococcal density in the nasopharynx, which has been shown to correlate positively with pneumococcal DNA detection in bronchoalveolar lavage samples, establishing the foundation for understanding how interventions affecting nasal colonization can influence pneumonia risk^10^.

Previous studies have reported nasopharyngeal colonization with pneumococcus primes alveolar macrophages (AMs) in the lung mucosa, amplifying their responsiveness to pneumococcus and other bacterial pathogens^10^. This priming effect can be persistent for three months and confers broad-spectrum increases in bacterial uptake against pathogens such as *Streptococcus pyogenes, Staphylococcus aureus* and *Escherichia coli*^10^. In the upper airway, LAIV administration has been shown to transiently increase pneumococcal carriage acquisition and bacterial density. Given that increased nasal density drives microaspiration into the lower airways, LAIV exposure can potenitally affect the amount of bacteria transcending into the lung mucosa^6^. LAIV-induced nasal inflammation also alters local immune responses, impairing neutrophil functions and monocyte recruitment^7^. Together, these observations raise the question on how prior LAIV exposure followed by pneumococcal infection reshape immune function (both innate and adaptive) in the distal lung and how that differs from TIV administration.

To systematically investigate how lower airway immune cells are affected by influenza immunization and subsequent pneumococcal infection, as in nasopharyngeal pneumococcal carriage, we used an Experimental Human Pneumococcal Challenge (EHPC) model. In this study design, participants received either LAIV intranasally, TIV intramuscularly, or no vaccine, followed by pneumococcal challenge 3 days later. Cross-sectional sampling of the lower airways was performed for a minimum of 30 days, and up to 3 months post immunization against influenza/pneumococcal challenge. Our experimental design of influenza vaccination preceding pneumococcal challenge directly models the clinically relevant scenario of whether different influenza vaccination strategies (LAIV as a proxy to an upperway contained influenza viral infection) alter host susceptibility to bacterial colonization acquisition, while focusing on the immunological changes caused by both vaccine, bacterial colonization and subsequent bacterial shredding to the distal lung.

We observed that LAIV vaccinees who established pneumococcal colonization after challenge had a higher frequency of influenza-specific CD4⁺ T cells in the lung mucosa and increased bacterial uptake capacity among alveolar macrophages (AMs). Single-cell RNA-seq confirmed AMs as the predominant cell type in BAL^11^, followed by monocyte-differentiating macrophages and T cells. It also revealed that vaccination differentially primed populations between four distinct AM gene programs linked to anti-microbial defense, antigen presentation, and phagocytosis, with changes in cytokines and T cell activation suggesting robust crosstalk between T cells and AMs. Together, these findings indicate that influenza vaccination reprograms the lung mucosal immune landscape through coordinated interactions between adaptive and innate cells. By showing that intramuscular and nasal vaccine platforms differentially shape macrophage gene modules and T cell activation in the lung mucosa, and that these effects are further modified by pneumococcal colonization, our work contextualizes vaccine responses within a dynamic host-microbe ecosystem. This perspective underscores how microbial exposures and vaccine platforms intersect to tune both protective immunity and immune homeostasis, providing a foundation for understanding why some individuals remain protected while others are at risk of post-influenza bacterial pneumonia, and pointing toward new strategies for next-generation respiratory vaccines and immunomodulatory interventions.

## Results

### Immunophenotyping and scRNAseq profiling of influenza vaccination and pneumococcal challenge clinical cohort

In a double-blinded, randomized clinical trial, we administered either tetravalent-inactivated influenza vaccine (TIV, intramuscular, blue) or live attenuated influenza vaccine (LAIV, nasal administration, red) to 117 healthy adults aged 18-48 years^12^. Vaccination preceded an intranasal challenge with live *S. pneumoniae* by three days, and pneumococcal colonization rates were similar in LAIV and TIV vaccinated subjects (25/55 [45.5%] vs. 24/62 [38.7%]; odds ratio [OR], 1.32; *P* = 0.46). Following the study follow-up period (Day 29 to Day 120 post challenge), BAL samples were collected from a subset of study participants who consented to research bronchoscopy. To assess and compare immune responses resulting from the influenza vaccines, we included healthy volunteers challenged with live *S. pneumoniae* without prior vaccination (Unvac & Chall., green) and healthy volunteers who did not receive any vaccination or bacterial challenge (Unvac & Not Chall., gray; **Fig 1A**). All BAL samples were collected within 30-120 days post-Spn bacterial challenge (**Supp Table 1**).

**Figure 1.**
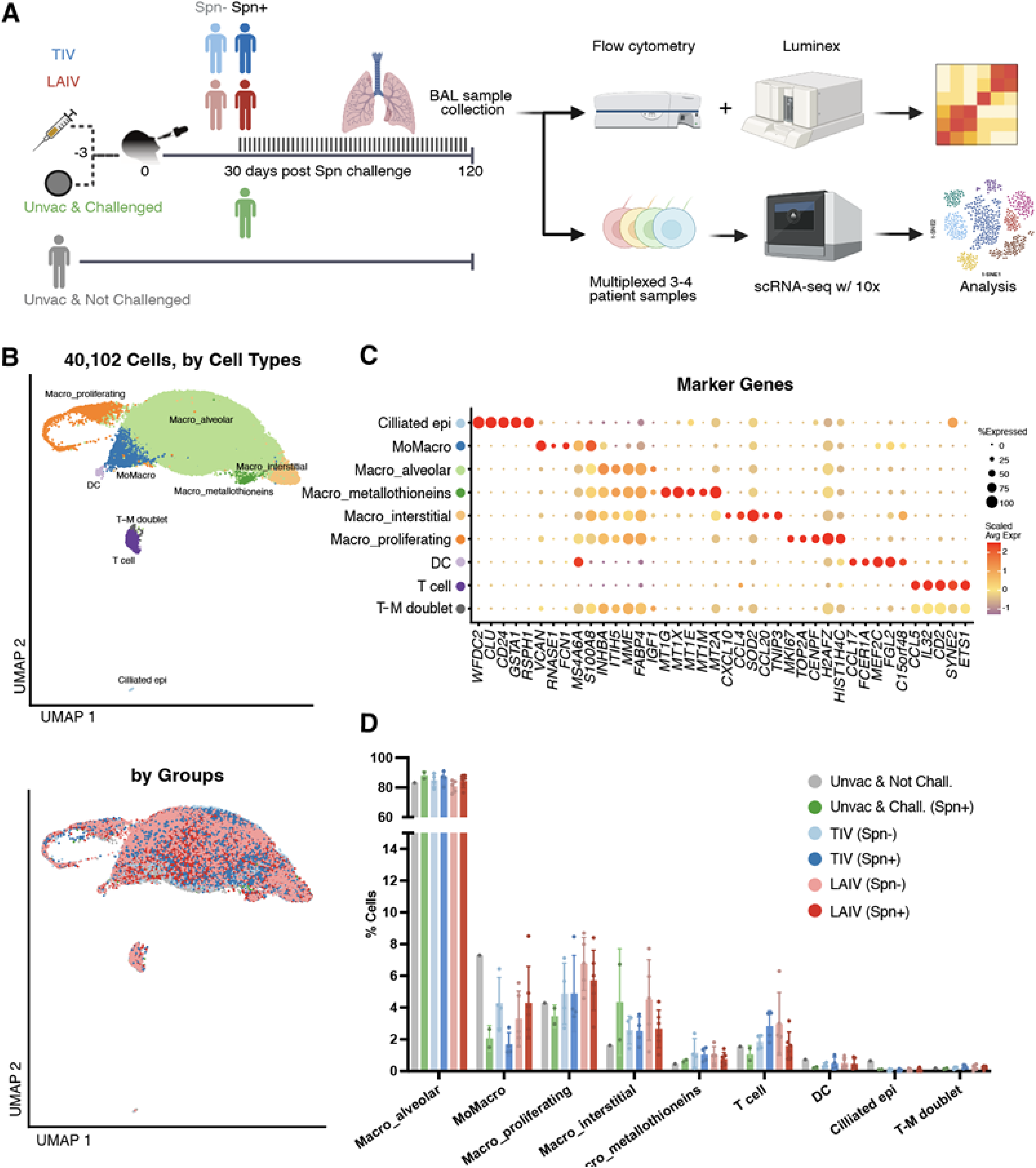
Immunophenotyping and scRNA-Seq profiling of influenza vaccination and pneumococcal challenge clinical cohort. **A**. Schematics for sample acquisition, processing, and analysis workflow of Bronchoalveolar Lavage (BAL) samples from an Experimental Human Pneumococcal Challenge (EHPC) model, of intramuscular inactivated (TIV) and nasal live attenuated (LAIV) influenza vaccines followed by *Streptococcus pneumoniae* (Spn) challenge. **B**. Uniform manifold approximation and projection (UMAP) visualization of 40,102 single cells colored by nine broad cell types (top) and by sample groups (bottom). **C**. Scaled RNA expression of top markers used to identify broad cell types. **D**. Difference in cell proportional composition across the 6 sample groups. No significant differences between groups were found using Fisher exact test or Dirichlet regression analysis. Ciliated epi, ciliated epithelial cells; MoMacro, transitioning monocytes; Macro_alveolar, alveolar macrophages; Macro_metallothioneins, metallothionein-expressing macrophages; Macro_interstitial, interstitial macrophages; Macro_proliferating, proliferating macrophages; T-M doublet, doublets formed by T cell and macrophage; DC, dendritic cells.

First, to evaluate global inflammatory responses, we performed measurement of 30 cytokines and chemokines by Luminex on the acellular component of these BAL samples (**Fig 1A, Supp Fig 1A**). As the BAL samples were performed 30-120 days post-challenge, this sampling captures the mucosal environment during the resolution and homeostatic phase rather than peak acute inflammation. Consistent with this timing, we did not observe broad, dramatic upregulation of inflammatory cytokines. Instead, we observed that colonized adults in the LAIV showed a decrease in inflammatory mediators G-CSF, IL13, IFNγ, IL2, IL2R, and Eotaxin (Wilcoxon rank-sum test; significant in LAIV group: p = 0.03, 0.01, 0.04, 0.04, 0.01, and 0.03 respectively), while colonized individuals in the TIV group showed reductions in IL7, IL10, and IL17 (Wilcoxon rank-sum test; significant in TIV group, p = 0.04, 0.04, 0.04, respectively).

Next, we evaluated whether there was any difference in humoral response in the lung mucosa. We found the levels of IgG to influenza antigens were 2.5-fold higher in TIV compared to LAIV vaccinees, whereas levels of anti-influenza IgA were 2.65-fold greater in the LAIV compared to TIV vaccinees (**Supp Fig 1B;** Kruskal-Wallis test, followed by Dunn’s test; p= 0.025 and p=0.041, respectively). We did not observe a significant correlation between the days post-challenge and the titers for IgG (p = 0.23 and 0.08 for LAIV and TIV, respectively) or IgA (p = 0.80 and 0.92 for LAIV and TIV, respectively) (**Supp Fig 1F**). This suggests that antibody levels are relatively stable within this window, and the differences observed between the vaccine groups are more likely reflective of the magnitude of the response driven by the respective vaccine platforms rather than sampling bias. In the case of post-challenge humoral responses to pneumococcus, we observed that Spn-colonized individuals had 2-fold higher levels of IgG in response to CPS-6B (*S. pneumoniae* 6B capsular polysaccharide of challenge strain) compared to non-colonized individuals (**Supp Fig 1C;** Kruskal-Wallis test, followed by Dunn’s test; p=0.0015). Pneumococcal carriage status did not confer any significant change to anti-influenza responses **(Supp Fig 1D)**. Similarly, influenza vaccine formulation did not alter antibody responses to Spn **(Supp Fig 1E).**

Next, to holistically examine the cellular and molecular factors associated with responses to vaccination and carriage, we performed scRNA-seq and flow cytometry on the cryopreserved BAL samples cells **(Fig 1A, Supp Fig 2A; Methods)** from 22 subjects. We obtained 40,102 high-quality cells across six conditions: unvaccinated/not challenged (n=1), unvaccinated/Spn+ (n=2), LAIV vaccinated/Spn- (n=5), LAIV vaccinated/Spn+ (n=6), TIV vaccinated/Spn- (n=4), and TIV vaccinated/Spn+ (n=4). Using dimensionality reduction and unsupervised clustering, we identified nine distinct major cell types **(Fig 1B)**.

These cell types encompass ciliated epithelial cells (Ciliated epi; *CD24, WFDC2*), myeloid cells including transitioning monocytes (MoMacro, *VCAN, FCN1*), alveolar macrophages (Macro_alveolar, *INHBA, FABP4*), metallothionein-expressing macrophages (Macro_metallothioneins, *MT1G, MT1X*), interstitial macrophages (Macro_interstitial, *CXCL10, CCL4, CCL20*), and proliferating macrophages (Macro_proliferating, *MKI67, TOP2A*), along with a small population of dendritic cells (DC, *FCER1A*) and lymphoid cells, as in T cells (T cell, *CD3E*) (**Fig 1C**). Initially, we examined relative frequency changes in cell types across groups. As anticipated, AMs were the predominant cell type in BAL samples, followed by T cells and DCs (**Fig 1D)**. As we did not observe significant differences in immune cell type proportions across groups, we decided to perform a systematic study of each major cell type (**Fig 1D, Supp Fig 2B**).

### Alveolar macrophages show transcriptomic differences across vaccination and carriage status

To elucidate cellular reprogramming as a function of vaccination and carriage, we focused on identifying the cell types with the greatest number of differentially expressed genes when comparing vaccination and carriage status. We systematically evaluated differential gene expression across vaccination and colonization groups within all recovered populations, including rarer subsets such as ciliated epithelial cells, DCs, and T cells. By performing differential gene expression analysis between vaccinated groups (TIV and LAIV) and non-vaccinated individuals in the context of pneumococcal carriage, we did not identify any significantly differentially expressed genes within these minor subsets, likely due to the limited recovery of these cells from BAL. Consequently, we chose not to emphasize these populations in the current analysis, as their limited numbers restrict their contribution to the central conclusions of this study. Instead, we focused our deep transcriptional profiling on alveolar macrophages (AMs), which displayed the largest and most robust transcriptomic shifts (**Fig 2A**), and their interactions with broader cell types such as DC and T cells which displayed fewer differentially expressed genes (DEGs). Genes specifically upregulated in AMs among all vaccinated subjects were associated with cell-cell adhesion (*IGFBP2, IGF*) and antigen presentation (*HLA, FCER1G*), and increased metabolic activity (glucose transporter, SLC2A3; **Fig 2B, 2C**; **Supp Table 2**). In addition, the myeloid inhibitory C-type lectin-like receptor (*CLEC12A*) was more highly expressed in AM from vaccinated subjects, consistent with previous findings reporting *CLEC12A* potentiates antiviral responses (**Fig 2B**)^13^. Similar gene enrichments were observed for interstitial and proliferating macrophages(**Supp Fig 3A**). No differentially expressed genes were detected in T cells and DC between vaccinated and unvaccinated groups, however, which could be due to limited recovery of these cells from BAL (averaged 39 T cells and 9 DC per volunteer).

**Figure 2.**
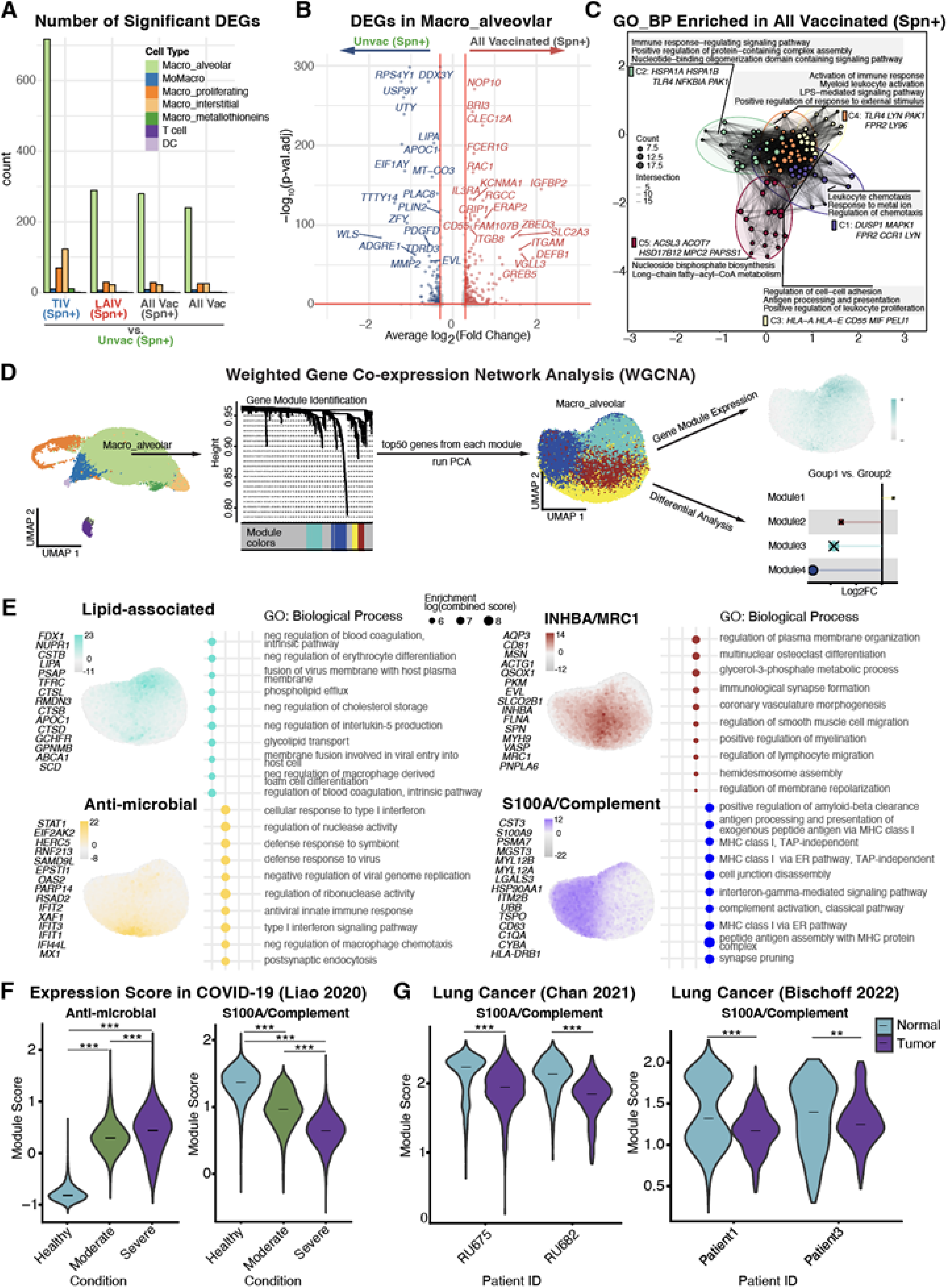
Alveolar macrophages exhibit distinct transcriptional programs shaped by vaccination and colonization. **A.** Number of significantly differentially expressed genes for each cell type by comparing vaccinated groups (TIV/LAIV) vs unvaccinated groups (Unvac) for individuals with pneumococcal carriage. **B**. Volcano plots of differentially expressed genes in AMs when comparing all vaccinated groups to the unvaccinated group in colonized individuals. Genes upregulated in the vaccinated group are labeled in red and genes upregulated in the unvaccinated group are labeled in blue. **C**. Network plot for the enriched gene sets from the differentially expressed genes of alveolar macrophages in the vaccinated groups. **D.** Schematic of weighted gene co-expression network analysis (WGCNA) workflow on alveolar macrophage. Top 50 genes from each identified module were used to run principal component analysis (PCA) for UMAP visualization (**Methods**). **E.** Top 15 genes and their enriched pathways for each gene module. **F.** “anti-microbial” and “S100A/Complement” module expression in the context of COVID-19^22^. **G**. “S100A/Complement” module expression in the contexts of lung cancer^23,24^. Significance levels: ns = not significant; *p < 0.05; **p < 0.01; ***p < 0.001.

To identify patterns in the AM transcriptional differences between vaccinated and unvaccinated adults, we performed Weighted Gene Co-expression Network Analysis (WGCNA)^14^. This method allowed us to identify consensus gene modules that represent major gene programs in this population, for which we could then evaluate associations with vaccination and colonization status. After determining the optimal soft-thresholding power by testing scale-free topology fit and constructing a signed co-expression network based on the topological overlap matrix, we identified four significantly correlated modules (**Fig 2D**). Subsequently, each cell was scored by its expression of the gene modules to enable visualization and downstream cell-cell interaction analysis (**Fig 2E**)

Within these four modules, we identified a) a “lipid-associated” module that displayed enrichments in biological processes linked to phospholipid efflux and glycolipid transport (*APOC1, ABCA1, CD36*) and b) an “INHBA/MRC1” module that featured genes related to phagocytosis (*MRC1, CD81, MYH9*), representing fundamental AM biological processes of relevance to the host immune response (**Fig 2E**). We also identified c) an “anti-microbial” module highly enriched in genes associated with antiviral response and type I interferon signaling pathway (*STAT1, OAS2, IFIT1, IFIT2, IFIT3, IFI44L, MX1*), whereas the fourth gene module (S100A/Complement) was highly enriched in complement activation (*C1QA*), antigen presentation (*FCER1G, PSMB3, PSMB7, CYBA, HLA-DRB*), and interferon-gamma signaling pathways (*IFI30* and genes related to class I and II MHC molecules) (**Fig 2E, Supp Table 3**). To better understand how these programs relate to known macrophage features, we further scored each cell using “classically activated” M1 and “alternatively activated” M2 macrophage polarization signature genes modules^15^. Across AMs, elevated expression of the “lipid-associated” and “INHBA/MRC1” modules was associated with higher M2-like signature expression as compared to the other modules (p.adj < 0.001, Wilcoxon rank-sum test, **Supp Fig 3B**).

To gain deeper insights into the drivers of each gene module within AMs, we conducted transcriptional factor (TF) inference analysis using DoRothea^16^ ^17^. This analysis revealed that upregulation of TFs was enriched in two modules: the “anti-microbial” and “S100A/Complement” modules. TFs associated with antiviral functions, including IRF1, IRF9, STAT1, STAT2, and STAT5, were upregulated in the “anti-microbial” module (**Supp Fig 3C**), whereas the “S100A/Complement” module was enriched for TFs associated with innate defense mechanisms and antigen recognition. Specifically, we inferred elevated activities for NF-KB, a key factor for pro-inflammatory responses, and STAT4, a TF crucial in responding to IL12/IL23 and type I interferon and essential for directing type 1 T-helper cell differentiation to combat pathogens^18^ (**Supp Fig 3C**).

As expected for scRNA-seq data, we detected low levels of most cytokine transcripts (**Supp Fig 3D**). Therefore, to infer potential downstream cytokine consequences of TF activity, we used DoRothea’s TF-target database (**Supp Fig 3C**). Prioritizing genes for cytokines measured at the protein level within the Luminex cytokine panel (**Supp Fig 1A**), we found evidence to suggest that both the “anti-microbial” and “S100A/Complement” modules have the potential to induce pro-inflammatory cytokines through activating STAT1 and IRF1 transcription factor activities, such as IL10, IL12A, IL12B, and CXCL10. The “S100A/Complement” module may induce other inflammatory cytokines by activating the NF-KB signaling pathway, such as CXCL8, CXCL9, TNF, and GM-CSF (CSF2), showing its possible involvement in early innate defense mechanisms, cell recruitment of neutrophils and peripheral blood monocytes to the lung mucosa, and adaptive immunity through antigen presentation ^19,20^. These transcriptomic findings provide a mechanistic basis for our direct cytokine measurements. The “S100A/Complement” module is enriched for transcription factors predicted to drive the production of pro-inflammatory cytokines (**Supp Fig 3C**). Therefore, the decreased levels of specific inflammatory cytokines observed via Luminex in colonized adults (**Supp Fig 1A**) serve as a protein-level validation of this suppressed AM “S100A/Complement” transcriptional program.

To ensure more global relevance of our AM gene modules across various disease contexts, we leveraged a publicly available reference comprising 12 scRNA-seq studies across multiple human lung diseases^21^. Through a comparative analysis of the expression scores of our gene modules alongside 20 gene programs (GEPs) documented in this reference, we identified a high correlation between our “anti-microbial” module and GEP9, as well as between the “S100A/Complement” module and GEP8 (**Supp Fig 3E**). Notably, their program GEP9, akin to our “anti-microbial” module, is associated with antiviral responses, and exhibits enrichment in severe patients with COVID-19 and in subjects with pulmonary fibrosis compared to healthy samples (**Fig 2F**). Conversely, in the same COVID dataset of the BAL samples from the reference^22^, we observed elevated expression of the “S100A/Complement” module in healthy individuals compared to patients with moderate or severe symptoms (**Fig 2F**). Furthermore, we evaluated the “S100A/Complement” module in two additional lung cancer datasets with paired normal-tumor samples^23,24^ to evaluate its importance in cancer contexts. In these, we also observed elevated levels of the “S100A/Complement” module in normal tissues, aligning with our hypothesis that this module plays a pivotal role in preserving lung environment homeostasis, regardless of whether the challenge stems from infectious diseases or cancer (**Fig 2G**).

To further identify the potential stimuli that prime the expression of each gene module in AM, we correlated the gene module expression with a set of downstream genes stimulated by 59 different ligands from a publicly available dataset of *in vitro* macrophage stimulation^25^ (**Supp Fig 3F).** Here, we found that the “anti-microbial” module was highly correlated with IFNB1-stimulated genes, consistent with its predicted function while the “S100A/Complement” module was associated with TNF and IFNγ cytokine stimulation.

### “Anti-microbial” and “S100A/Complement” gene programs are associated with phagocytosis capability and driven by vaccine and colonization status

As AMs play a key role in clearing bacteria in the lower airways, we assessed their capacity to take up bacteria *in vitro* using an opsonophagocytic assay. We observed a consistent decrease in opsonophagocytic activity over time across groups and within groups (**Fig 3A**: Pearson R = - 0.49, P = 0.014, **Supp Fig 4A:** LAIV: p = 0.048; TIV: p = 0.2). We observed that AMs in LAIV/Spn+ group had the highest opsonophagocytic activity amongst groups, particularly compared to non-vaccinated non-colonized individuals (median 73% uptake in LAIV/Spn+ vs 58% in Unvac/Spn-; Kruskal-Wallis test, followed by Dunn’s test for multiple comparison correction; p=0.043) and non-colonized LAIV recipients (median 73% in LAIV/Spn+ vs 54.8% in LAIV/Spn-, Kruskal-Wallis test, followed by Dunn’s test for multiple comparison correction; p=0.0575; **Fig 3B**).

**Figure 3.**
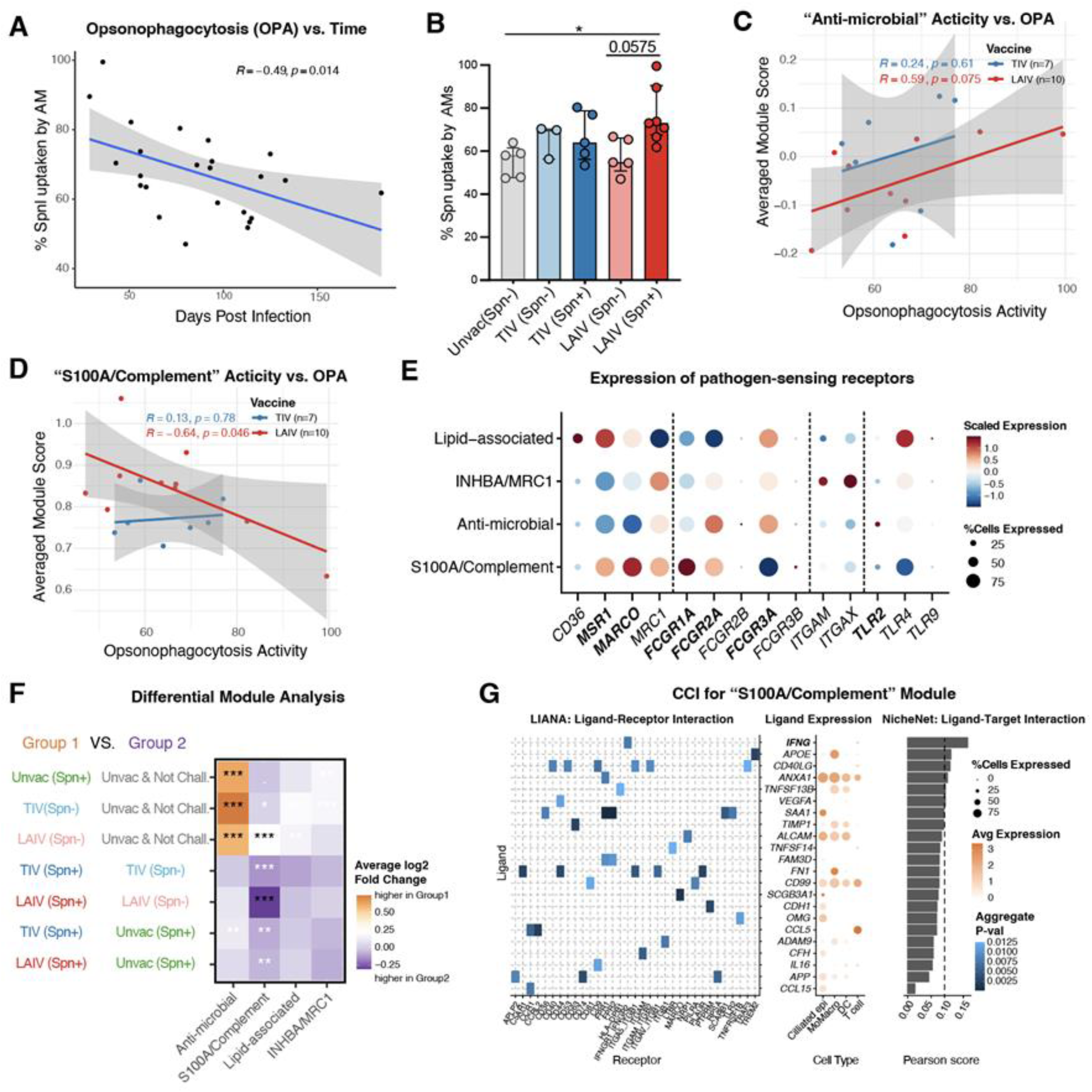
“Anti-microbial” and “S100A/Complement” gene programs are associated with phagocytosis capability and driven by vaccine and colonization status. **A.** Pearson correlation of opsonophagocytic activity of AMs and days post pneumococcal challenge (p = 0.014). **B.** Opsonophagocytic activity of AMs measured by the percentage of pneumococcal uptake in the BAL samples. Significance was first assessed using Kruskal-Wallis test, followed by Dunn’s test for multiple comparison correction. The corrected p-value between LAIV (Spn-) and LAIV (Spn+) is 0.0575, at the borderline of significance. *p < 0.05; **C-D.** Correlation of the functional opsonophagocytic activity of AMs in each individual and their averaged expression of **(C)** the “anti-microbial” module (TIV: p = 0.61; LAIV: p = 0.075) and (**D**) the “S100A/Complement” module (TIV: 0.78; LAIV: 0.046). **E.** Expression of pathogen-sensing receptors across AM modules. Receptors are grouped by functional class (dashed dividers): scavenger receptors (CD36, MSR1, MARCO, MRC1), Fc gamma receptors (FCGR1A, FCGR2A, FCGR2B, FCGR3A, FCGR3B), integrins (ITGAM, ITGAX), and Toll-like receptors (TLR2, TLR4, TLR9). **F**. Differentially expressed modules by comparing different vaccination and colonization status. Orange color indicates higher module expression in Group 1 and Purple color indicates higher expression in Group 2. P values are labeled in black color if the absolute average log2 fold change is above 0.2. **G**. Cell-cell interaction (CCI) analysis showing top ligands that can act on macrophage and induce the “S100A/Complement” module. Left panel: the aggregate p-value from LIANA representing the interaction strength for top ligands (y-axis) and associated receptors (x-axis); middle panel: dotplot showing ligand expression in sender cell types; right panel: NicheNet pearson score indicating the strength of the ligands that can induce the “S100A/Complement” module. High Pearson score (>=0.1) suggests strong evidence that the ligand is active in driving the “S100A/Complement” module expression. Significance levels: ns = not significant; *p < 0.05; **p < 0.01; ***p < 0.001.

To understand the functional consequences of the AM gene programs we identified above, we averaged the expression scores of each module for individual adults and correlated them with their corresponding AM opsonophagocytic ability that we experimentally measured. We found that adults with higher “anti-microbial” module scores had augmented AM opsonophagocytic ability (Pearson R = 0.59, p = 0.075 in the LAIV group), indicating the activation state of the alveolar macrophages to fight against microbial infection **(Fig 3C)**. In contrast, the “S100A/Complement” module was negatively correlated with opsonophagocytic ability (Pearson R = -0.64, p = 0.046 in the LAIV group), reflecting a potential scavenger role and the activated state phase of the alveolar macrophage (**Fig 3D**). To understand the functional association of these two AM modules, we looked for expression of pathogen-sensing receptors (**Fig 3E**). The “anti-microbial” module is marked by elevated *FCGR2A*, *FCGR3A*, and *TLR2* expression with reduced *MARCO* and *MSR1*, which is a configuration favoring antibody-mediated phagocytosis of Gram-positive bacteria^26–28^, consistent with its positive correlation with opsonophagocytic activity (**Fig 3C**). Conversely, the “S100A/Complement” module has high expression of *MARCO* and *FCGR1A*, receptors that are associated with ability to trigger NF-KB-coupled signaling and cytokine production^29,30^, consistent with our TF inference (NF-KB, STAT4) and pro-inflammatory cytokine secretion (**Supp Fig 3C**), suggestive of a sensing-and-signaling phenotype specialized for inflammatory coordination rather than opsonized particle uptake.

To explicitly uncover the distinct and interacting effects of pneumococcal colonization and influenza vaccination on alveolar macrophage states, we evaluated differential gene module usage by leveraging eigengene module expression across our cohorts (**Fig 3F**). Our data suggest context-dependent regulation of the “Anti-microbial” and “S100A/Complement” module. Specifically, the “anti-microbial module” is enriched in non-vaccinated subjects colonized by pneumococcus and in vaccinated subjects without pneumococcal colonization, indicating its dual functions in anti-bacterial and antiviral responses, respectively. This is consistent with its positive correlation with opsonophagocytic activity and an increased antibody-dependent effector activity, as reflected by the higher expression of the Fcγ receptors FCGR2A and FCGR3A. Further comparison between subjects receiving TIV and LAIV revealed that TIV triggered a slightly higher expression of the “anti-microbial” module in AMs than LAIV (**Supp Fig 4B**) This is consistent with TIV inducing substantially higher influenza-specific IgG responses than LAIV, both systemically and in the pulmonary mucosa^31^,which may drive greater Fc receptor-mediated macrophage activation and partially account for this observation. On the other hand, in both vaccination groups, subjects not colonized by pneumococcus showed significantly higher expression of the “S100A/Complement” module than colonized subjects. This transcriptomic difference is corroborated by our direct cytokine measurements: colonized adults had lower levels of inflammatory cytokines in BAL (**Supp Fig 1A**). Because the “S100A/Complement” module is enriched for transcription factors predicted to drive pro-inflammatory cytokine production, its downregulation in colonized subjects offers a plausible cellular explanation for reduced cytokine levels. Furthermore, vaccination exerted a distinct effect on macrophage state in the context of bacterial exposure: among pneumococcus-colonized subjects, both AMs in the LAIV and TIV groups expressed the “S100A/Complement” module at lower levels than non-vaccinated controls. This suggests that prior vaccination, combined with bacterial colonization, further shifts lung homeostasis toward a state with fewer AMs expressing the “S100A/Complement” module.

While the cross-sectional design of this human challenge model precludes definitive confirmation of causality, these structured module comparisons provide robust evidence of coordinated functional state shifts in the lung mucosa. We frame these findings as indicative of dynamic macrophage reprogramming events, specifically regarding the balance between the “anti-microbial” and “S100A/Complement” axes, that warrant further targeted mechanistic validation in future longitudinal or functional studies.

Since the expression of the “S100A/Complement” module displayed major changes across comparison groups, we asked what ligands in the tissue could modulate this gene module by performing cell-cell interaction analysis (CCI; **Methods**). We found that IFNγ is the top ligand, highly expressed in T cells, that is predicted to act on AMs’ surface receptors and induce the expression of the “S100A/Complement” gene module (**Fig 3G**). These results lead us to investigate other major cell types in our dataset further.

### Nasal vaccination and colonization recruit more monocyte-differentiating macrophages and alter antigen processing capacity

Next, we looked into other myeloid cell types in the dataset. Even though most proportional changes were not significant when contrasting vaccination and carriage statuses, we observe an increase of monocyte-differentiating macrophages (MoMacro) for colonized individuals in the LAIV group compared to the TIV group (**Supp Fig 4C,** LAIV Spn+ vs. TIV Spn+: p = 0.043, Wilcoxon test). Furthermore, a subset of colonized individuals who received LAIV showed a higher pneumococcal bacterial density compared to those who received TIV (**Supp Fig 4D),** suggesting that nasal vaccination may indirectly enhance monocyte recruitment to the lower airways in response to increased bacterial load. Looking at differentially expressed genes in MoMacro subpopulations, in colonized individuals, we found that the LAIV group upregulated *ERAP2* and *RPS4Y1* while downregulating HLA related genes compared to the TIV group (**Supp Fig 4E**). Given that *ERAP2* (Endoplasmic Reticulum Aminopeptidase 2) is critical for peptide processing in macrophages^32,33^ and HLA molecules are required for antigen presentation, these transcriptional changes suggest that this recruited monocyte-differentiating macrophages retain antigen processing capacity but may be limited in their ability to present antigens compared to professional APCs like DCs. Notably, The combination of increased MoMacro abundance and upregulated *RPS4Y1* expression (previously associated with asthma susceptibility^34^) may reflect a disturbance in local immune homeostasis, consistent with clinical reports linking LAIV administration in children under two years of age to an increased risk of wheezing^35,36^.

### Both intramuscular and nasal immunization routes activate T cells in the lung mucosa

As T cells have been shown to play a pivotal role in driving immune response to vaccination, we investigated the effects of vaccination route and carriage status on T cell function in the lung mucosa. Subclustering, the T cell population from the scRNA-seq data and using the automated cell type annotation tool *CellTypist*, we were able to define 4 major T cell subtypes: CD4 T cells (CD40LG, LTB), CD8 T cells (CD8A, CD8B), γδT cells (TRDC) and regulatory T cells (FOXP3, CTLA4) (**Fig 4 A-B**).

**Figure 4.**
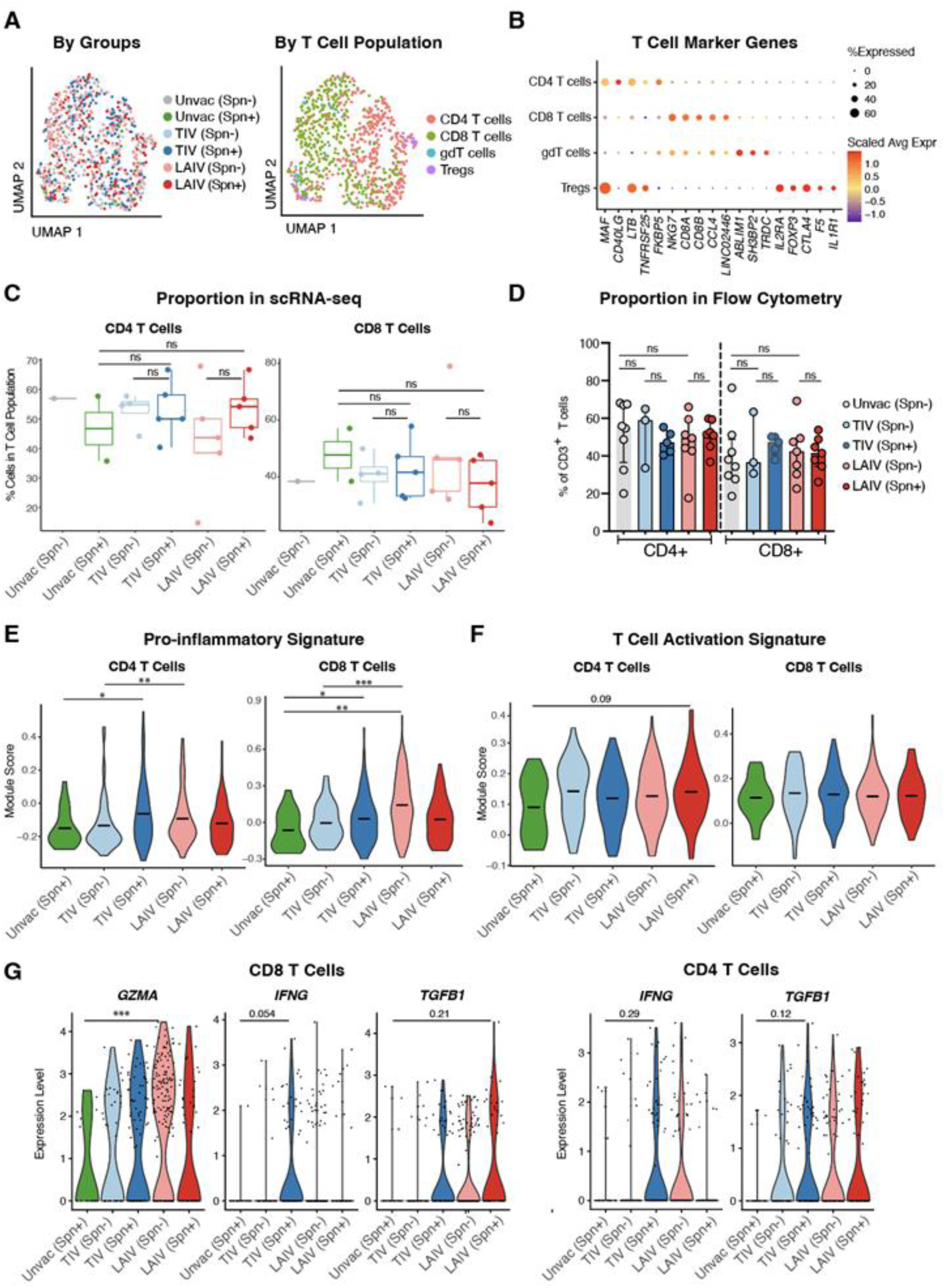
scRNAseq analysis of T cells reveals activation of CD4 T cells regardless of the vaccination type. **A**. UMAP visualization of 832 T cells colored by sample groups (left) and T cell subtypes (right). **B**. Scaled RNA expression top differentially expressed genes in each assigned T cell subtypes. **C**. Difference in T cell proportional composition across the 6 sample groups. No significant differences between groups were found using Fisher exact test or Dirichlet regression analysis. **D**. Composition of T cell subsets in BAL samples in unvaccinated (n = 8), TIV/Spn-(n=3), TIV/Spn+ (n=5), LAIV/Spn- (n=7) and LAIV/Spn+ (n=7) individuals. **E.** Expression scores of a pro-inflammatory signature^37^ and **F.** activation signature^38,39^ on CD4+ (left) and CD8+ (right) T cells. **G.** Single-cell level expressions for the transcripts encoding granzyme A (*GZMA*), interferon gamma (*IFNG*), and TGF beta (*TGFB1*) in CD8+ (left) and CD4+ (right) T cells. Unvac, unvaccinated; TIV, tetravalent-inactivated influenza vaccine; LAIV, live attenuated influenza vaccine; Spn, *Streptococcus pneumoniae*; BAL, bronchoalveolar lavage. γδ, gamma-delta T cells; DN, double negative T cells. Nominal significance levels: ns = not significant; *p < 0.05; **p < 0.01; ***p < 0.001.

Influenza vaccination with either TIV or LAIV was not associated with substantial changes in the abundance of CD4 or CD8 T cells when comparing vaccination routes and colonization status (**Fig 4C; Methods**). This same pattern was also observed by flow cytometry (**Fig 4D, Supp Fig 4F**). Given the limited recovery of T cells in our BAL samples, we present our single-cell T cell analysis as a higher-level, signature-based assessment rather than a deeply resolved subset analysis. Therefore, we focused on comparing overarching T cell states by calculating and contrasting functional signature scores using the single-cell RNA expression data - including signatures of pro-inflammatory^37^, T cell activation/GO:0042110, type I interferon-mediated signaling pathway/GO:0060337, type II interferon-mediated signaling pathway/GO:0060333^38,39^—rather than attempting to resolve granular subpopulation differences (**Supp Fig 5A-B**). We found that, in non-colonized individuals, LAIV induced higher pro-inflammatory activity in both CD4 and CD8 T cells (**Fig 4E**, LAIV/Spn-vs. TIV/Spn-, CD4: nominal p = 0.0116; CD8: nominal p = 0.0009, adjusted p = 0.006, Kolmogorov-Smirnov test). In addition, in colonized individuals, TIV induced both CD4 and CD8 T cells to a more pro-inflammatory state compared to the unvaccinated group (**Fig 4E**, Kolmogorov-Smirnov test; TIV/Spn+ vs. Unvac/Spn+, CD4: nominal p = 0.048; CD8: nominal p = 0.049). We observed a similar trend for a T cell activation signature in CD4 cells, even though the statistical test was not significant, likely due to the limited sample size (**Fig 4F**). Next, we examined key genes important for T cell function, such as interferon gamma (IFNG), granzyme A (GZMA) and transforming growth factor-beta (TGFB1). In single-cell expression data, we identified that both immunizations drive CD8 T cells to upregulate *GZMA* (**Fig 4G**, Kolmogorov-Smirnov test; in CD8 GZMA expression, LAIV/Spn-vs Unvac/Spn+: nominal p=0.0008, adjusted p = 0.007). Although shifts in IFNG and TFGB1 expression were not statistically significant, we did observe a similar trend of upregulation in vaccination groups for both CD4 and CD8 T cells.

### Live attenuated influenza vaccine with pneumococcus co-infection enhances antigen-specific CD4+ T cell responses in the lower airway

To further evaluate T cell shifts, we leveraged flow cytometry to identify antigen-specific T cell populations. Among influenza-specific T cells, there was a trend towards increased TNF^+^IFNγ^+^ CD4^+^, but not CD8^+^, T cell frequency in the lung mucosa of the LAIV vaccinated group (global CD4: median 0.25 in LAIV vs 0.065 in TIV recipients) (**Fig 5A**). However, when the vaccinated individuals were further stratified by pneumococcal colonization status, an increased frequency of influenza-specific TNF^+^IFNγ^+^ CD4^+^ T cells (both global and TRM CD4^+^), but not CD8^+^, was detected in the LAIV/Spn+ compared to the non-colonized groups (global CD4^+^:median 0.7285 in LAIV/Spn+ vs 0.0 in LAIV/Spn- and TIV/Spn-; p=0.007 and p=0.006, respectively; TRM CD4^+^:median 0.47 in LAIV/Spn+ vs 0.0 in LAIV/Spn- and TIV/Spn-; p=0.009 and p=0.016, respectively; Kruskal-Wallis test, followed by Dunn’s test; **Fig 5B**). In agreement with previous studies^2,10^, the frequency of Spn-specific CD4^+^ T cells (global and TRM CD4^+^) was greater in the lung mucosa of Spn-colonized individuals (median 0.19 of global CD4^+^ and 0.32 of TRM CD4^+^ in the Spn+ group vs 0.0 of both CD4+ and TRM CD4+ in the Spn-group, p=0.006 and p=0.029, respectively; Kruskal-Wallis test, followed by Dunn’s test; **Fig 5C**). Vaccination with either TIV or LAIV did not have any substantial effect on T cell responses to pneumococcus (**Fig 5D).**

**Figure 5.**
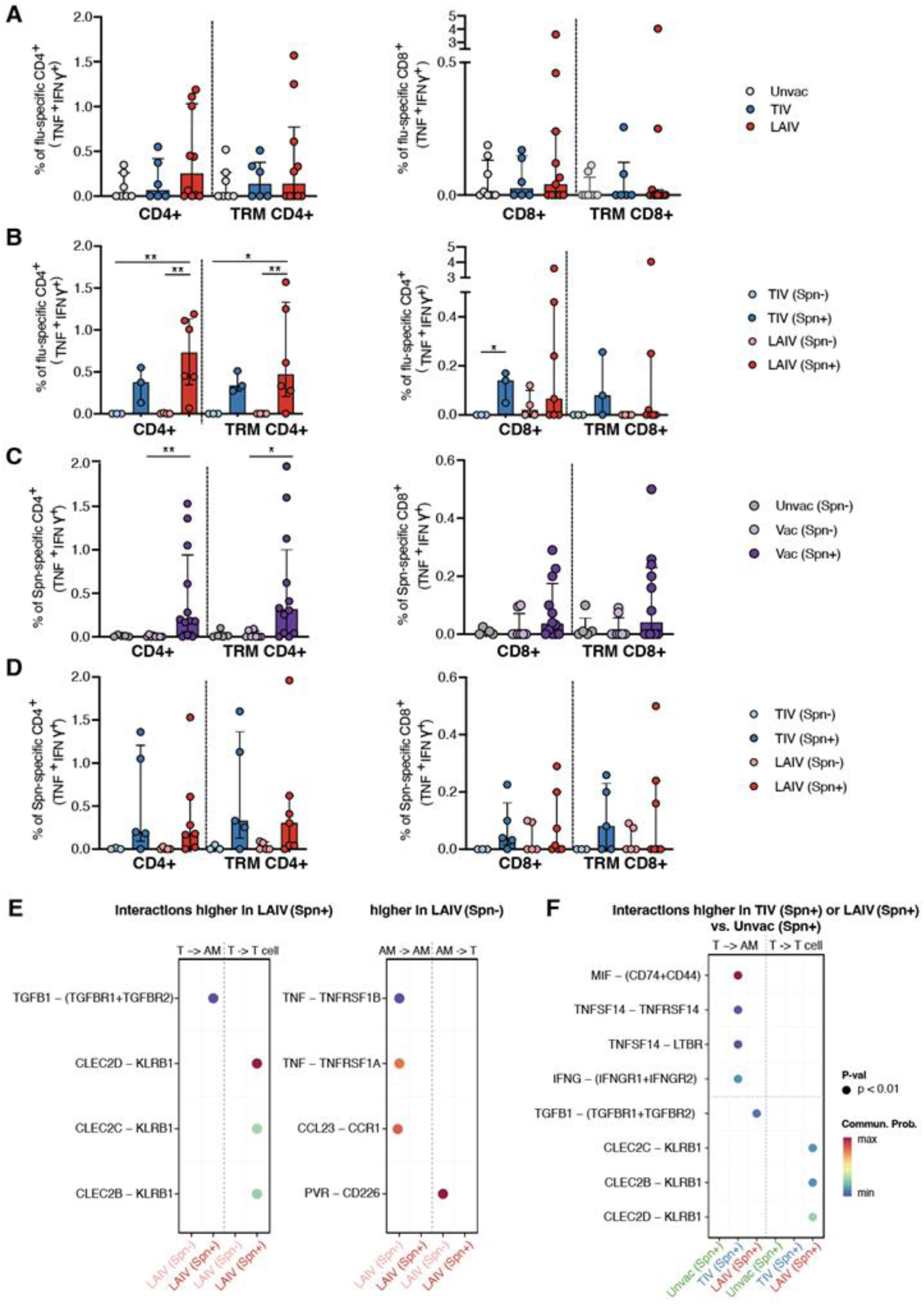
Live attenuated influenza vaccine with pneumococcus co-infection enhances antigen-specific CD4+ T cell responses in the lower airway. **A**. Frequency of flu-specific CD4+ (left) and CD8+ (right) T cells in the global and TRM population in unvaccinated (n = 8), TIV (n=6) and LAIV (n=11) groups. **B**. Frequency of Spn-specific CD4^+^ (left) and CD8^+^ (right) T cells in the global and TRM population in TIV/Spn- (n=3), TIV/Spn+ (n=3), LAIV/Spn- (n=4) and LAIV/Spn+ (n=7) groups. **C**. Frequency of spn-specific CD4^+^ (left) and CD8^+^ (right) T cells in the global and TRM population in unvaccinated/Spn- (n=5), vaccinated/Spn- (n= 8) and vaccinated/Spn+ (n = 12) groups. **D**. Frequency of spn-specific CD4^+^ (left) and CD8^+^ (right) T cells in the global and TRM population in TIV/Spn- (n=3), TIV/Spn+ (n=5), LAIV/Spn- (n=5) and LAIV/Spn+ (n=7) group. Significance was assessed using Kruskal-Wallis test, followed by Dunn’s test for multiple comparison correction. Significance level: *p < 0.05; **p < 0.01. **E–F.** Differential T cell-AM signaling across colonization and vaccination conditions, inferred by CellChat. Dot color indicates the communication probability (blue, low; red, high) and dot presence indicates statistical significance (p < 0.01, Wilcoxon rank-sum test); **E.** Comparison of colonized (Spn+) versus uncolonized (Spn-) LAIV recipients. **F.** Comparison of colonized TIV and LAIV recipients against unvaccinated colonized (Unvac/Spn+) controls.

### Inferred T cell-AM communication shift toward a regulatory network with vaccination and colonization

To connect the antigen-specific T cell responses with the alveolar macrophage states defined earlier, we used CellChat to ask whether the T cell-macrophage communication network was reshaped by colonization and vaccine route (**Methods**). We first isolated the effect of colonization within LAIV recipients, where T cell numbers were sufficient for inference (**Fig 5E**). The inferred network reorganized in a biologically coherent direction: Spn-colonized individuals displayed a T cell-centric regulatory network dominated by TGFB and inhibitory CLEC2-KLRB1 signaling, whereas uncolonized individuals displayed an AM-centric inflammatory and surveillance network, characterized by TNF secretion, CCL23-mediated recruitment, and PVR-CD226 cytotoxic stimulation of T cells. Within colonized individuals, both vaccine groups showed enhanced T cell-driven signaling absent in unvaccinated controls, but the mode differed by route (**Fig 5F**): TIV recipients signaled through effector-type ligands (MIF, TNFSF14, IFNG), while LAIV recipients engaged macrophages via TGFB and self-regulated through CLEC2-KLRB1, mirroring their weaker pro-inflammatory T cell signature (**Fig 4E**). Even where IFNγ-driven T cell-to-AM signaling was inferred (most clearly in TIV recipients), the IFNγ-responsive “S100A/Complement” module remained reduced in colonized recipients of both vaccines (**Fig 3F**), indicating that its suppression tracks with colonization-associated regulation rather than the mere availability of inferred IFNγ input. Both axes therefore converge on the LAIV/Spn+ group as the most regulatory state, the same group showing the strongest antigen-specific CD4+ TRM response and the most pronounced decrease of the “S100A/Complement” AM module expression.

## Discussion

In this study, we leveraged an Experimental Human Pneumococcal Challenge model to dissect how influenza vaccination and pneumococcal colonization jointly shape immunity in the lower respiratory tract. By integrating single-cell transcriptomics with functional immune assays, we provide a detailed view of how adaptive and innate immune cells are reprogrammed not in isolation, but through coordinated crosstalk.

Our data shows that LAIV recipients who became colonized with *Streptococcus pneumoniae* exhibited the strongest influenza-specific CD4⁺ T cell responses in the lung mucosa (Fig 5B), accompanied by enhanced opsonophagocytic activity of alveolar macrophages (Fig 3B). This suggests that bacterial colonization can act as a natural adjuvant, amplifying vaccine-induced mucosal immunity^2,3,6,10^. Strikingly, these benefits occurred despite LAIV replication being largely confined to the upper airway^7,12,31^, reinforcing the concept that events in the nasopharynx can shape immune states in the distal lung. Prior work has shown that pneumococcal density in the nose correlates with microaspiration into the lower airway, seeding bacterial signals that prime AMs^10^. Our findings suggest that this seeding, combined with vaccine-induced T cell activation, creates a synergistic environment that enhances antiviral defense.

Yet, this synergy may be a double-edged sword. AMs from LAIV/Spn⁺ individuals displayed both the high “anti-microbial” activity and the lowest expression of the “S100A/Complement” module, which has been linked to antigen presentation and homeostatic regulation^21–24^. This pattern suggests that dual viral-bacterial stimulation may enhance immediate clearance capacity while perturbing macrophage programs that normally control their inflammation state. Mechanistically, the IFNγ-iNOS axis likely contributes to this state: IFNγ is an inferred upstream regulator of the “S100A/Complement” module and, through iNOS, can enhance anti-microbial activity via reactive nitrogen intermediates^40^. IFNγ also promotes the production of reactive oxygen species (ROS), another critical effector mechanism by which AMs kill intracellular and extracellular pathogens^41^. Notably,this module is suppressed in colonized vaccinees despite their elevated T cell activation, and IFNγ protein is itself reduced in their BAL (Supp Fig 1A) even as T cell IFNγ transcript only trends upward (Fig 4G). Rather than a simple loss of IFNγ-iNOS/ROS drive, this pattern is most consistent with active immunoregulation of the inflammatory program (a T cell-macrophage regulatory circuit we resolve below) that restrains the module while immediate anti-microbial capacity is retained. Because sampling is cross-sectional, we cannot fully separate ongoing regulation from a feedback state following earlier activation.

Beyond AMs, our dataset also uncovered reprogramming in monocyte-derived macrophages. Colonized LAIV recipients upregulated ERAP2 (implicated in peptide trimming) while downregulating HLA class II genes, suggesting these recruited cells may process antigens but present them inefficiently^42^. This imbalance could weaken local antigen presentation, limiting the quality of T cell priming despite the influx of monocytes. To our knowledge, this represents a novel observation of how nasal vaccination and bacterial carriage jointly skew macrophage differentiation. Together, these observations align with prior murine studies showing that influenza exposure can transiently reprogram the lung macrophage compartment through recruitment of monocyte-derived macrophages with heightened innate responsiveness. In particular, Aegerter *et al* ^43^ demonstrated that post-influenza monocyte-derived alveolar macrophages enhance anti-bacterial defense, with this imprint driven by macrophage origin rather than stable reprogramming of resident AMs. Our human data suggest that influenza vaccination and pneumococcal colonization engage a related macrophage differentiation axis in the distal lung.

The adaptive arm of immunity also reflected vaccine-specific signatures. Whereas TIV elicited stronger influenza-specific IgG in the BAL, LAIV preferentially boosted IgA, in line with systemic versus mucosal routes of delivery^31,44^. This compartmentalization underscores that different vaccine strategies excel at different arms of protection: TIV for systemic humoral immunity, LAIV for mucosal antibodies and T cells. Both vaccines drove lung CD4⁺ and CD8⁺ T cells toward more pro-inflammatory and activated states, including expression of *IFNG* and *GZMA*^17,18^. However, as captured by our Luminex data (Supp Fig 1A), these transcriptional changes were accompanied by an overall dampening of broader inflammatory signaling networks in the BAL fluid of colonized individuals. This reflects a complex interplay between viral vaccination and bacterial colonization: the lower airway can sustain localized, targeted anti-viral and anti-bacterial cellular defenses without sustaining widespread, potentially damaging mucosal inflammation^7,31^. This localized precision is a highly desirable feature for vaccines intended to safely enhance mucosal immunity.

These observations raise the question of how the lower airway sustains heightened antigen-specific defense without tipping into damaging inflammation. Our cell-cell interaction analysis points to a candidate hypothesis (Fig 5E-F). Within colonized LAIV recipients, the inferred T cell-AM network was dominated by regulatory TGFB and inhibitory CLEC2-KLRB1 signaling (absent in the unvaccinated individuals), whereas uncolonized recipients showed an inflammatory, AM-centric signaling. We propose that LAIV combined with pneumococcal colonization may establish a coordinated T cell-macrophage circuit in which heightened antigen-specific T cell activity is paired with active immunoregulatory signaling, accounting for both the localized precision and the restrained “S100A/Complement” program noted above. This interpretation is based on ligand-receptor inference in a small cohort with limited T cell recovery, and key ligands such as TGFB1 were not individually significant at the transcript level (Fig 4G); it should therefore be regarded as a hypothesis for future testing. Recent work showing that vaccine-elicited CD4 T cells can durably imprint alveolar macrophages and sustain antiviral programs in the lung offers a plausible basis for this crosstalk and a framework for testing its directionality directly^10,45,46^.

Finally, the AM modules we defined showed strong concordance with transcriptional programs in public BAL datasets from COVID-19 and lung cancer^22–24^. This convergence suggests that the “anti-microbial” versus “homeostatic” balance we describe represents a conserved axis of lung immunity that can be perturbed across diverse disease contexts. Thus, our findings extend beyond influenza, and pneumococcus, highlighting shared macrophage-T cell circuits that may be targeted in other inflammatory or infectious settings^21,47^.

Our study is limited by its small sample size due to the intrinsic complexities and the cross-sectional nature of these types of challenges and BAL sampling. Nevertheless, the reproducibility of our macrophage modules across public datasets and their functional correlations with phagocytosis provide confidence in their relevance. Future work should include longitudinal sampling to track how these macrophage states evolve over time, experimental models to probe causality of the IFNγ-iNOS/ROS axis, enrichment of less abundant populations such as DCs and B cells and integration with the lung epithelial compartments to capture the full mucosal network^48–50^

In summary, our findings reveal that mucosal influenza vaccination and pneumococcal colonization converge to reprogram lung immunity through dynamic interactions between T cells and macrophages. While this interplay enhances antigen-specific T cell responses and immediate anti-microbial activity, it also rebalances alveolar macrophages away from the homeostatic “S100A/Complement” program and, in recruited monocyte-derived macrophages, may limit antigen-presentation capacity. These observations offer important translational insights for future respiratory vaccine strategies. Our data support the concept that the immunological context of the respiratory mucosa at the time of vaccination profoundly shapes the quality of downstream lung immune responses. The association between pneumococcal colonization and enhanced influenza-specific CD4⁺ T cell responses in LAIV recipients suggests that pre-existing mucosal microbial exposures may function as natural modifiers, or even adjuvant-like signals, of vaccine-induced immunity.

Additionally, the differential effects of LAIV and TIV observed here further underscore that the route of vaccine delivery matters. While both TIV and LAIV altered lower airway immunity, they engaged partially distinct immune programs. LAIV was more strongly associated with driving mucosal cellular responses in the setting of colonization, highlighting the potential advantages of mucosal vaccination approaches for deliberately inducing robust, antigen-specific T cells while the local immune environment.

Ultimately, our work positions the lung mucosa as an active ecosystem in which viral vaccination and bacterial colonization jointly determine whether host defenses are protective, dysregulated, or both. This suggests that future vaccine evaluation must consider the broader host-microbe ecosystem rather than vaccine formulation alone. Variables such as colonization status, baseline airway immune tone, and resident macrophage states likely contribute to interpersonal variability in vaccine efficacy. Recognizing this layered crosstalk is essential for designing next-generation respiratory vaccines and immunomodulatory interventions aimed at enhancing protective mucosal immunity while simultaneously preserving delicate tissue homeostasis.

## Limitations of the study

This study has several limitations. First, the sample size was constrained by the ethical and logistical demands of bronchoalveolar lavage sampling in a controlled human infection model, limiting power for analyses of less abundant immune populations. For rarer populations, such as ciliated cells, DCs, and T cells, the limited cell counts precluded deeply resolved subset analyses or the robust detection of differentially expressed genes. Therefore, we restricted our single-cell evaluations of these populations to higher-level, signature-based assessments and limited our primary comparisons to the groups with sufficient sample numbers, focusing our central conclusions on the more abundantly recovered macrophage populations.. Findings related to the smaller subgroups should be interpreted with appropriate caution, serving primarily as hypothesis-generating observations. Second, airway sampling was cross-sectional and performed 30-120 days after vaccination and pneumococcal challenge. Due to the invasive nature and ethical constraints of performing research bronchoscopies in healthy volunteers, it was not possible to collect longitudinal BAL samples from the same individuals over time. This restricts precise resolution of the temporal dynamics underlying macrophage and T cell state transitions, as well as the temporal kinetics of the mucosal antibody responses. Third, we note that two participants in the TIV (Spn–) group showed evidence of an asymptomatic viral infection during their screening visit. However, this occurred several months prior to bronchoscopy (a week before vaccination and pneumococcal challenge), and neither individual had an ongoing respiratory viral infection on the day of BAL sampling. Given this temporal separation, a sustained effect on interferon-driven programs at the time of sampling is highly unlikely. Consistently, these individuals did not present as outliers in our single-cell transcriptomic analyses, and their inclusion did not skew group-level patterns. Rather, their inclusion reflects the natural immunological variability expected in human cohort studies. Additionally, although our integrative analyses implicate IFNγ-associated pathways in macrophage-T cell crosstalk, causal relationships cannot be directly tested in humans and are inferred from convergent transcriptional, cytokine, functional, and ligand-receptor analyses. Finally, the use of a single pneumococcal serotype in healthy adults may limit generalizability to other host populations or clinical settings. Despite these constraints, the controlled human challenge design provides rare in vivo insight into how vaccination and bacterial colonization jointly shape lung immunity.

## Data Availability

Lead contact
Further information and requests for resources and reagents should be directed to and will be fulfilled by the lead contact, Elena Mitsi.
Materials availability
This study did not generate new reagents.
Data and code availability
The raw files for the datasets generated in this study are available at (Currently in the process to be submitted to GEO). This paper does not report original code. Any additional information required to reanalyze the data reported in this paper is available from the lead contact upon request.

## Resource availability

### Lead contact

Further information and requests for resources and reagents should be directed to and will be fulfilled by the lead contact, Elena Mitsi.

### Materials availability

This study did not generate new reagents.

### Data and code availability

The raw files for the datasets generated in this study are available at (Currently in the process to be submitted to GEO). This paper does not report original code. Any additional information required to reanalyze the data reported in this paper is available from the lead contact upon request.

## Acknowledgments

We would also like to thank all the volunteers for their participation in the clinical trial and the clinical team members for helping to collect the samples.

This work was funded by grants from the Bill & Melinda Gates Foundation: OPP1117728 awarded to DMF and a GH-VAP (OPP1202327/INV-006897) awarded to AKS and EM. The funder had no role in study design, data collection and analysis, decision to publish, or preparation of the manuscript. S.T. was supported in part by a Pew Latin American Fellowship Program in Biomedical Sciences.

## Authors Contribution

E.Y.T. performed wet-laboratory experiments, analyzed the data, and wrote the manuscript, S.T. performed wet-laboratory experiments, analyzed the data, and wrote the manuscript, D.D.R performed wet-laboratory experiments, analyzed the data, and wrote the manuscript, J.Reine., SPJ, OA P.C.S. performed wet-laboratory experiments and analysed data. J.Rylance recruited study participants and collected clinical samples. A.K.S, D.M.F. and E.M. conceived the study, provided intellectual input, supervised the study, and provided insights into data outputs.

## Supplementary Table legends

**Supp Table 1.** Demographic and clinical characteristics of healthy adult volunteers included in this influenza vaccination and pneumococcal challenge study.

**Supp Table 2.** Differentially expressed genes in each broad cell type by comparing vaccinated unvaccinated individuals (related to Figure 2B).

**Supp Table 3.** Alveolar macrophage gene modules identified by WGCNA (related to Figure 2E).

## Methods

### Study Design: the BAL sample collection

Adult volunteers aged 18 to 50 years were enrolled in the parent LAIV clinical trial study (REC 14/NW/1460), registered on EudraCT (number 2014-004634-26) and ISRCTN (number 16993271). The trial was co-sponsored by the Royal Liverpool University Hospital and the Liverpool School of Tropical Medicine. Exclusion criteria included a prior history of influenza or pneumococcal vaccination, clinically confirmed pneumococcal disease in the preceding 2 years, pregnancy, close contact with individuals at increased risk for pneumococcal disease (children under 5, immunosuppressed people, and elderly people), recent febrile illness, current or recent use of antibiotics, or immune-modulating medication. Volunteers received either LAIV (Fluenz Tetra, n=53), or TIV (Fluarix Tetra, n=62), were challenged with pneumococcal serotype 6B (80,000 CFU per nostril) 3 days after the influenza vaccination and were followed up for 27days post challenge. Pneumococcal colonization was detected by classical microbiology methods, and individuals were defined as Spn-colonized, if any nasal wash culture following experimental challenge grew Spn serotype 6B (Spn6B).

22 BAL samples were collected from a subset of study participants after the completion of follow-up visits using flexible bronchoscopy. All Spn-colonized individuals received three doses of amoxicillin on Day 27 (end of the clinical trial), and BAL samples were obtained between 30 to 120 days after the intranasal pneumococcal challenge: LAIV vaccinated/Spn- (n=5), LAIV vaccinated/Spn+ (n=6), TIV vaccinated/Spn- (n=4), and TIV vaccinated/Spn+ (n=4). Additional BAL samples from a) non-vaccinated/non-challenged individuals (n=1) or non-vaccinated/ Spn+ individuals (n=2) were used as control groups, collected under other study protocols (REC no. 18/NW/0481).

### Flow Cytometry Assays

Non-adherent lower airway cells were counted and seeded at 1 × 10^5^ cells in 96-well plate in medium with RPMI medium, containing FBS (10% heat inactivated, Thermo Fisher Scientific) and antibiotic mixture (Penicillin-Streptomycin-Neomycin, Sigma). Cells were stimulated with 1.2 μg/mL influenza antigens (TIV, 2016/2017) or 5μg/mL heat-inactivated Spn6B or left unstimulated as negative control and incubated for 2 hours at 37°C, 5% CO2. Then, 1000× diluted GolgiPlug (BD Biosciences) was added, and cells were incubated for an additional 16 hours.

The following day, cells were harvested, washed with PBS and stained with Pacific Blue viability dye (LIVE/DEAD Fixable Dead Cell stain kit, Thermo Fisher Scientific), followed by anti-human conjugated antibody cocktail targeting surface proteins. Antibodies used in the present study include: CD3 APC-Cy7 (clone SK7, BD), CD4 PerCP-Cy5.5 (clone SK3, Biolegend), CD8 PE-Dazzle (clone CF596, Biolegend), CD103 BV605 (clone Ber-ACT8, Biolegend), CD69 BV650 (clone FN50, Biolegend) and TCRgd PE-Cy7 (clone 11F2, BD). After fixation and permeabilization, the cells were stained with intracellular markers IFNγ-PE (clone 4SB3, Biolegend) and TNFα-APC (clone Mab11, Biolegend). After 30 minutes, samples were washed with cold PBS and acquired on a BD LSRII flow cytometer using FACSDIVA v.9.0. FMOs (fluorescence minus one values) and unstimulated samples were used to determine gates applied across samples. Data were analyzed using FlowJo v.10.10.

### ELISA for anti-influenza IgG and IgA levels and anti-CPS6B IgG in BAL

Levels of IgG and IgA to influenza and IgG to 6B capsular polysaccharide (CPS6B) were quantified in the BAL fluid samples using ELISA, as previously described ^31,44^.

#### For Influenza

Briefly, 96-well plates (Nunc maxisorp) were coated with 100 μL 0.2 μg/mL TIV in PBS and left overnight at room temperature. For detection of IgG and IgA, a 1:5000 of anti-human IgG (Sigma, A9544) and 1:4000 of anti-human IgA (MilliporeSigma, A9669) was made in 0.1% BSA and 100 μL added to each well after washing and incubated at room temperature for 1 hour. Next, plates were washed, and 100 μL p-Nitrophenyl Phosphate (MilliporeSigma) was added to the wells.

#### For CPS6B

Briefly, 96-well plates were coated with 5 μg/mL of purified CPS6B (Statens Serum Institut). Anti-CPS6B IgG was detected by goat anti-human IgG (1/5000; Fc-specific) alkaline phosphatase (Sigma, A9544). Optical density was measured at 405 nm using FLUOstar Omega plate reader (BMG Labtech).

The OD of each well was measured at 405 nm using a FLUOstar Omega ELISA microplate reader (BMG Labtech), the average blank corrected value was calculated for each sample, and the data analyzed using Omega Analysis (BMG)

### Luminex Analysis of BAL Supernatant

The acellular BAL fluid was collected after centrifugation of the whole BAL sample (400 g for 10 min at 4°C), divided to 1ml aliquots and stored at −80°C until analysis. On the day of the analysis, samples were concentrated 10× (1 ml of BAL supernatant concentrated to 100μl using rotary vacuum concentrator RVC2-18), following acquisition using a 30-plex magnetic Luminex cytokine kit (ThermoFisher) and analyzed on a LX200 with xPonent3.1 software following manufacturer’s instructions. Samples were analyzed in duplicates, and BAL samples with a coefficient of variation >50% were excluded.

### AM Opsonophagocytic Assay

AM opsonophagocytic capacity was evaluated as previously described^10^. Briefly, live Spn serotype 6B were opsonized with human intravenous immunoglobulin (IVIG, Gamunex; Grifols, Inc.) at 37°C for 15 minutes. An opsonized bacterial strain, baby rabbit complement (Mast Group), and isolated AMs were incubated at 37°C for 1 hour. Following incubation, 10 μl of reaction mixture was plated, in triplicate, onto blood agar (Oxoid) and incubated at 37°C, 5% CO2 overnight. Colony-forming units from cell supernatants were counted the following day. Multiplicity of infection used was 1:100.Due to the lack of readout of AM lysosome acidification, the term pneumococcal uptake rather than pneumococcal killing was used.

### Sample multiplexing and scRNA-seq

BAL samples were thawed and washed with RPMI medium FBS (10% heat-inactivated, Thermo Fisher Scientific) and antibiotic mixture (Penicillin-Streptomycin-Neomycin) at 37°C. After centrifugation of the sample (350 g for 5 min at 4°C), the supernatant was removed and replaced with PBS. Cells were counted and diluted to a concentration of 1 million cells per milliliter with 1x PBS as the diluent. Cells were stained with one of TotalSeq™-A0251 Antibody anti-human Hashtag 1 (BioLegend 394601), TotalSeq™-A0252 Antibody anti-human Hashtag 2 (BioLegend 394603) or TotalSeq™-A0253 Antibody anti-human Hashtag 3 (BioLegend 394605) according to manufacturer’s instructions. Cells from three BAL samples, each undergoing staining with a unique Hashtag antibody, were counted, and an equal number of cells from each sample were mixed together. These mixtures were further diluted using 1x PBS to the appropriate concentration for use with 10x Genomics Single-Cell 3′ Library Kit v3.1 NextGem (10× Genomics) protocols.

### scRNA-seq library preparation

Single-cell suspensions were loaded onto the 10x Chromium controller using the 10x Genomics Single-Cell 3′ Library Kit v3.1 NextGem (10× Genomics) according to the manufacturer’s instructions. In summary, cell and bead emulsions were generated, followed by reverse transcription, cDNA amplification, split into gene expression cDNA and antibody hash cDNA, fragmentation, and ligation with adaptors followed by sample index PCR. The resulting libraries were quality-checked by Qubit and Bioanalyzer, pooled, and sequenced using NextSeq2000 P3 100 cycles (Illumina) and Novaseq 6000 S4 (Illumina)

### Pre-processing of scRNA-Seq data

Raw sequencing data were processed using the CellRanger software (version 7.2.0). Gene expression reads were aligned to a custom reference human genome (GRCh38) and antibody hash reads to a reference list with the corresponding BioLegend barcodes. The resulting unique molecular identifier (UMI) count matrices were imported into R (version 4.1.2) and processed with the R package Seurat (version 4.0.1)^51^. Independent assays for RNA and Cell Hashing were added to the Seurat Object. Cell Hashing counts were normalized with a centered log-ratio transformation. Cells were then demultiplexed using the HTODemux Seurat function, and cells exhibiting a single unique identity were retained for downstream analysis.

### Cell type identification and proportion analysis

Cell type identification was performed using an integrative approach that combined reference-based label transfer with unsupervised marker discovery. Initial broad cell types were annotated using CellTypist automated annotation models along with “Cells_Lung_Airway” as reference^48–50^. To refine these annotations, we employed Seurat’s FindAllMarkers function with default parameters, applying the Wilcoxon Rank Sum test and setting a log2 fold-change threshold greater than 0.25, with markers required to be detected in at least 25% of cells within each cluster. This approach enabled the identification of top differentially expressed genes (DEGs) that distinguished both major cell types and their respective subtypes. For more focused analysis, macrophage and monocyte populations, as well as T cell populations, were subset from the dataset and subjected to further subclustering to resolve biologically relevant subpopulations unique to specific conditions. T cell subtype annotations were assigned by transferring labels from published single-cell RNA sequencing datasets of T cell samples^47^, utilizing Seurat’s anchor-based integration workflow^52^.

### Cell type proportion analysis

Cell type proportion analysis was conducted on the entire dataset as well as on these cellular subtypes, quantifying changes in cell type abundance and expression profiles across experimental groups. To account for the dependencies between cell subset proportions in scRNA-seq data, we used both Fisher exact tests and a Dirichlet multinomial regression analysis as complementary approaches to look for shifts in cell frequencies across conditions. We performed Fisher exact tests using the number of cells from each cell subset between two conditions to test whether a cell type was enriched in one condition (with Benjamini-Hochberg multiple testing correction). To account for the proportions of all other cell types in comparison, we also used the Dirichlet multinomial regression model from the DirichletReg R package^53^ and calculated the P values associated with abundance shifts (with Benjamini-Hochberg multiple testing correction).

### Differential gene analysis

For the identification of condition-specific gene interaction networks we performed differential gene co-expression analysis using the Model-based Analysis of Single-cell Transcriptomics (MAST) framework^54^. This approach enables us to identify coordinated changes that are connected to an experimental condition but may be indiscernible from noise when traditional differential expression analyses are leveraged. These genes provide better insights into the rewiring of gene regulatory networks in response to different vaccination and colonization status. MAST implements a hurdle model, combining a logistic regression component for gene detection frequency and a Gaussian linear model for normalized expression levels in detected genes. To control for technical confounders such as batch effects and inter-sample variability, sample identity was included as a latent variable in the model. Genes were considered differentially co-expressed if they exhibited a significant interaction term with a false discovery rate (FDR) below 0.05 and an absolute log2 fold-change greater than 0.5 in covariance strength. The utilization of these criteria increases our confidence pertaining to the identification of gene pairs whose coordinated expression patterns differ between conditions, thereby revealing context-dependent regulatory interactions that may underlie phenotypic differences.

### WGCNA and differential gene module analysis

Weighted Gene Co-expression Network Analysis (WGCNA) was implemented using the WGCNA R package to systematically identify conserved and condition-specific gene modules associated with vaccination and colonization status by characterizing higher-order transcriptional coordination patterns that reflect shared regulatory mechanisms or functional pathways^14^. Briefly, we created a gene-gene correlation adjacency matrix used to determine the soft power threshold that would minimize noise in the correlation matrix most effectively allowing for the retention of only strong connections. Modules of highly co-expressed genes were identified through topological overlap matrix-based hierarchical clustering to balance biological specificity with statistical robustness. Module eigengenes, representing the first principal components of each gene cluster, were correlated to condition groups aimed at identifying clinically relevant networks. Hub genes within significant modules were defined by intramodular connectivity measures (kME >0.25) and were included in an overrepresentation analysis aimed at identifying the enrichment of relevant immune response pathways. This network-based approach enabled the identification of vaccination- or colonization-associated gene modules that remained undetected in single-gene analyses but whose quantification provides insights into unique and shifted network rewiring present in our conditions when compared to baseline transcriptional variation.

To generate UMAP visualization for the AM population and demonstrate the distribution of module expression, we top 50 genes from each module and run PCA to perform dimension reduction. We also tested using top 100 genes from each module and the overall UMAP representation does not change significantly (data not shown).

### TF and downstream target inference

To infer transcription factor (TF) activity and identify downstream regulatory networks, we utilized DoRothEA, a curated resource of TF-target gene interactions with confidence-ranked regulons^16,55^. TF activities were calculated using the VIPER algorithm, which estimates regulatory protein activity by analyzing the enrichment of target gene expression patterns within each regulon. This method accounts for the pleiotropic effects of TFs by considering both positively and negatively regulated targets, with activity scores reflecting the net regulatory influence on downstream pathways. Downstream target networks were reconstructed by integrating significant TF activities (p-value <0.01) with their associated regulons, prioritizing targets showing coordinated expression changes across conditions. This dual-pronged approach was utilized to identify both master regulators driving phenotypic differences and their effector gene networks. The resulting TF-target networks were functionally annotated using overrepresentation analysis of Gene Ontology terms and pathway databases, with particular focus on immune response pathways.

### Signature correlation analysis

In order to statistically assess differential expression of coordinated expression patterns across gene modules and transcriptional regulatory networks, signature correlation analysis was implemented using Pearson correlation coefficients. The analysis focused on identifying conserved correlation structures that persist across biological replicates while detecting condition-specific covariation patterns indicative of condition-induced immune reprogramming. Pairwise Pearson correlations between signatures were calculated and correlation significance was assessed with false discovery rate (FDR) correction for multiple hypothesis testing (Benjamini-Hochberg procedure, FDR <0.05).

### Cell-cell interaction analysis

#### LIANA and NicheNet

Cell-cell communication was inferred using a two-step strategy combining LIANA^56^ for ligand-receptor prioritization with NicheNet^57^ for downstream target-gene activity prediction. Ligand–receptor interactions across all cell types were scored with LIANA, which integrates multiple scoring methods (using default five methods) into a consensus aggregate rank; The aggregated results were filtered to interactions originating from candidate sender populations (ciliated epithelial cells, MoMacro, DC, and T cells) and directed at the four AM modules, retaining only interactions with an aggregate rank p value below 0.05. For each receiver AM module, the top 100 ranked interactions were carried forward to NicheNet. Ligand activity was predicted with the prior-built NicheNet ligand-target matrix, ligand-receptor network, and weighted networks, with the geneset of interest defined as the WGCNA module gene set for each AM module. Candidate ligands were ranked by Pearson correlation between predicted and observed target-gene regulation. Using this approach, IFNγ emerged as the top-ranked ligand predicted to drive the “S100A/Complement” macrophage module. Unless otherwise stated, all other parameters were left at package defaults.

#### CellChat

To compare cell-cell interactions across vaccination/colonization status, we used CellChat with the human ligand-receptor database^58^. For each cell group, the analysis was restricted to genes in the ligand-receptor database, and over-expressed genes and interactions were identified before computing communication probabilities. Communications supported by fewer than 10 cells in a given cell group were removed (filterCommunication, min.cells = 10). Group-specific CellChat objects were then merged (mergeCellChat) for pairwise and three-way comparisons, with a focus on T cell-macrophage crosstalk. Using this approach, we compared colonized versus uncolonized LAIV recipients, and compared TIV- and LAIV-vaccinated colonized recipients against unvaccinated colonized controls. Unless otherwise specified, all parameters were left at CellChat defaults.

## Supplementary Figure legends

**Supp Figure 1.**
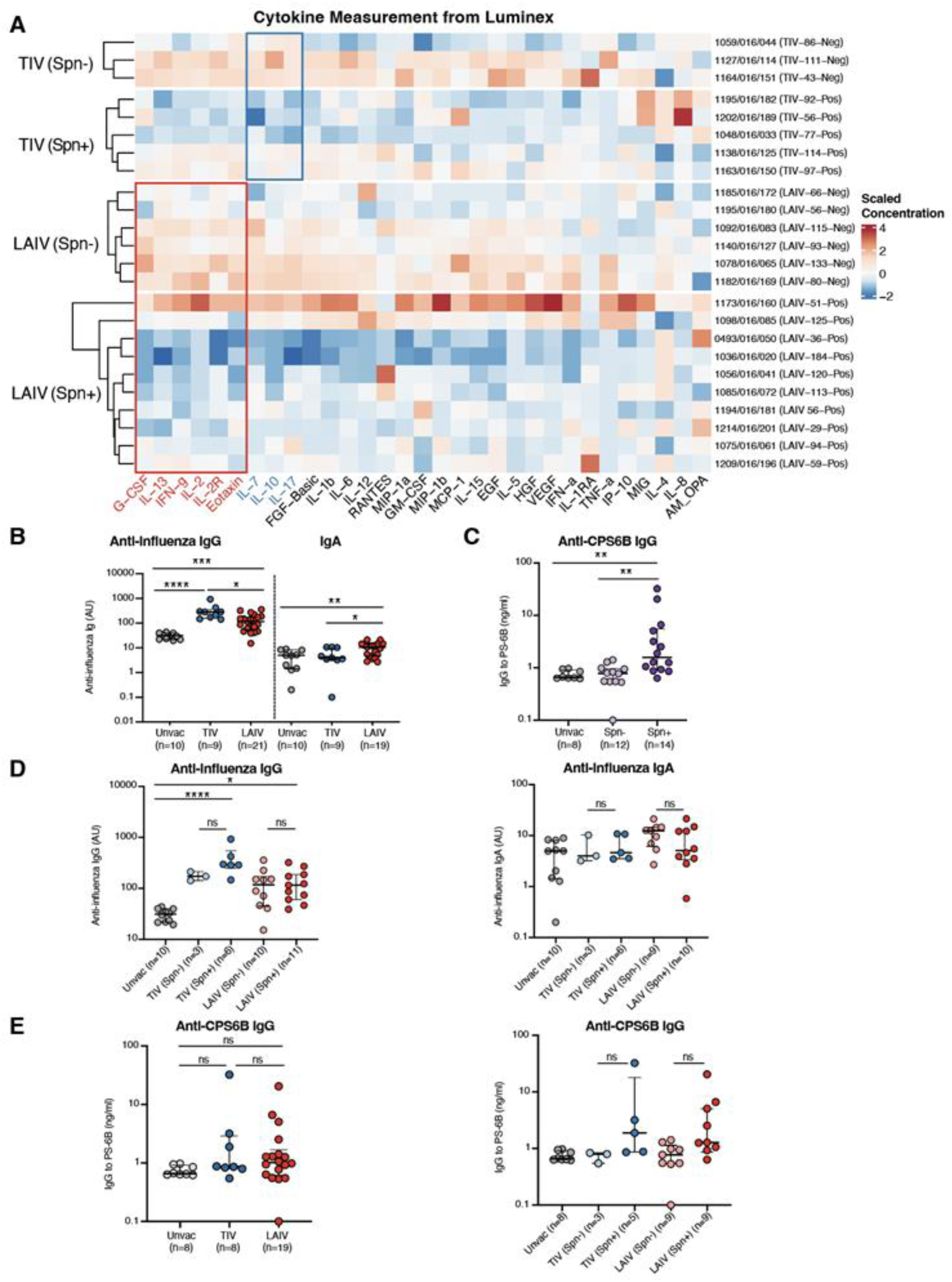
**A.** Luminex measure of Cytokine levels from the BAL samples. **B.** Levels of anti-influenza IgG and IgA by vaccination status. **C.** Anti-CPS6B IgG levels by carriage status. **D.** Levels of anti-influenza IgG and IgA by vaccination and carriage status. **E.** Anti-CPS6B IgG levels by vaccination and carriage status. Significance levels: ns = not significant; *p < 0.05; **p < 0.01; ***p < 0.001.

**Supp Figure 2.**
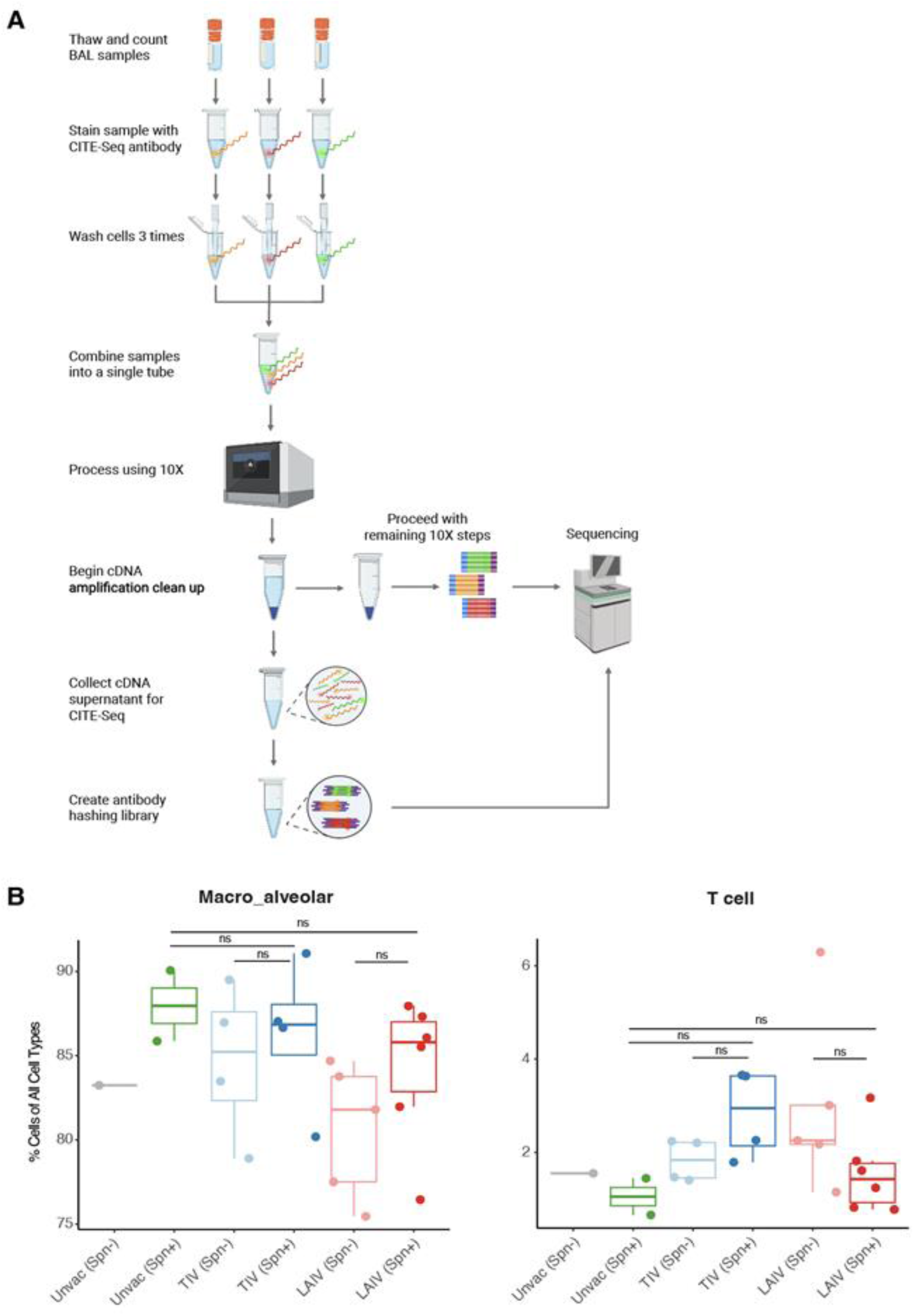
**A**. Schematic of experiment approach used for single-cell RNAseq. **B.** Difference in T cell and Macrophages proportional composition across the 6 sample groups. No significant differences between groups were found using Fisher exact test or Dirichlet regression analysis. Significance levels: ns = not significant.

**Supp Figure 3.**
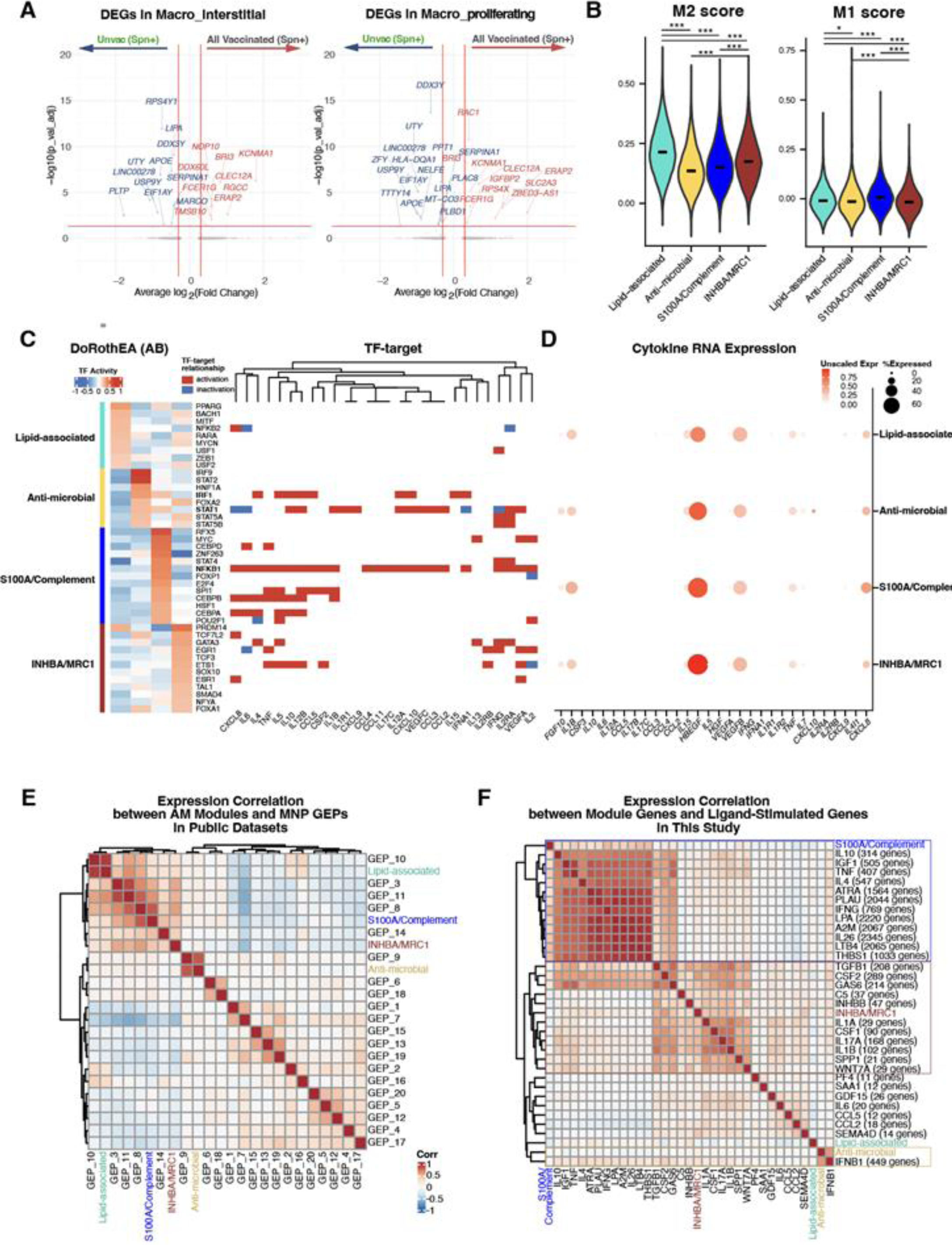
**A.** Volcano plots of differentially expressed genes (DEGs) in interstitial macrophages (Macro_interstitial) and proliferating macrophages (Macro_proliferating) when comparing all vaccinated groups to the unvaccinated group in colonized individuals. **B.** M1 and M2 signature score on the 4 AM modules. **C.** (left) DoRothEA transcription factor (TF) inference for each gene module. Inflammation-related TF activities were bolded. Only TF-target interactions with a high confidence level were selected for this analysis. (right) TF to target relationship for all 30 cytokines measured by Luminex for the same BAL samples. Only cytokines that are captured by RNA sequencing are shown. **D.** Cytokine expression in each macrophage population. **E.** Gene expression correlation of our identified AM modules and 20 gene programs defined in Peter et al’s reference for mononuclear phagocytes across lung diseases^21^. **F.** Gene expression correlation of our identified AM modules and ligand-stimulated gene sets derived from in vitro macrophage stimulation experiment16. Gene sets with strong correlations were boxed. Significance levels: ns = not significant; *p < 0.05; **p < 0.01; ***p < 0.001.

**Supp Figure 4.**
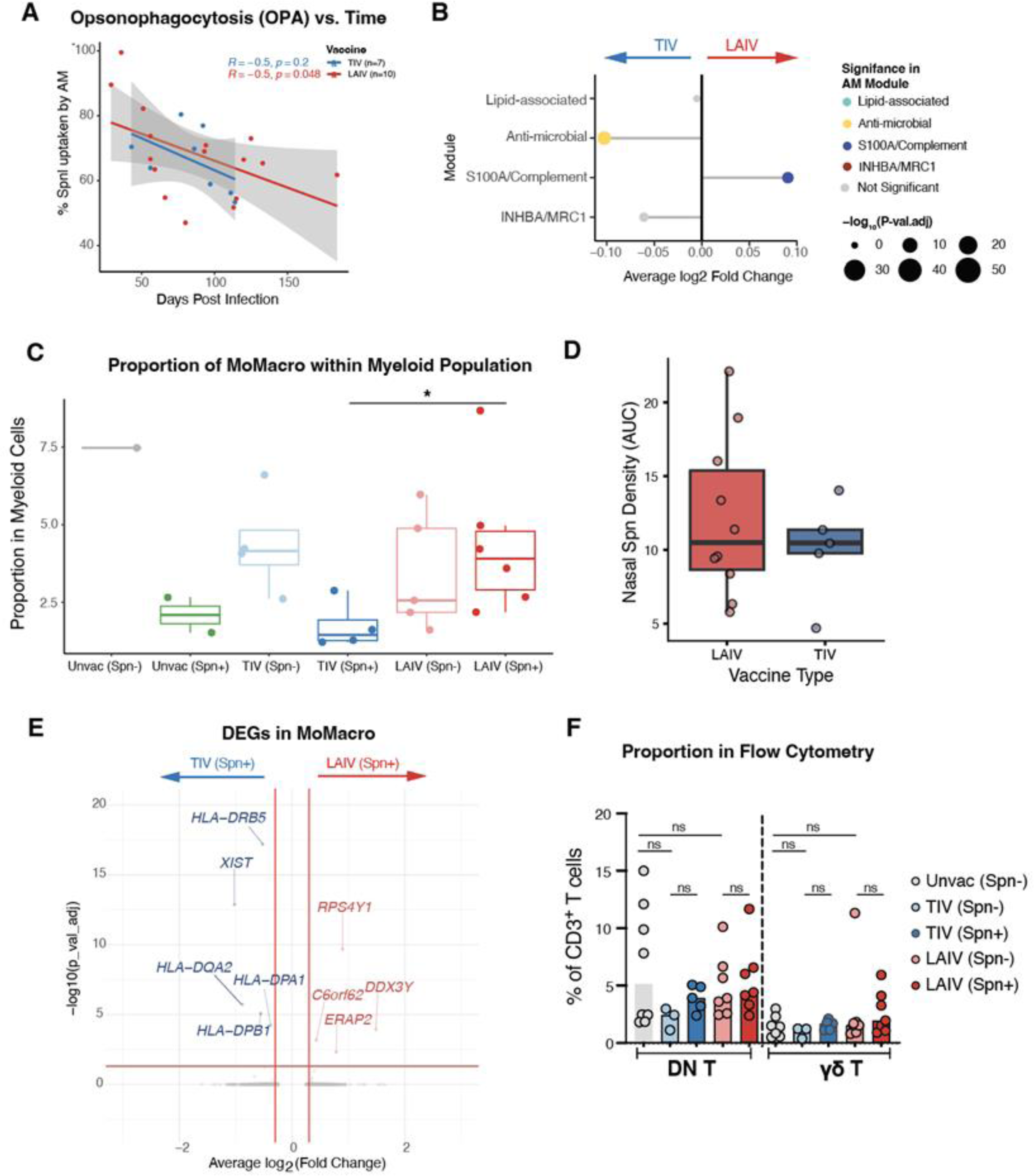
**A.** Correlation of Opsonophagocytic activity of AMs (AM.OPA) from the BAL samples measured by Luminex per vaccine type with day post-infection (LAIV: p = 0.048; TIV: p = 0.2). **B.** Differentially expressed modules by comparing TIV and LAIV groups. **C**. Difference in MoMacro proportional composition within myeloid lineage across the 6 sample groups. **D.** Calculated Area Under the Curve (AUC) for pneumococcal density during colonization for LAIV and TIV. **E.** Volcano plots of differentially expressed genes in MoMacro when comparing LAIV group to TIV group in colonized individuals. **F.** Composition of additional T cell subsets in BAL samples in unvaccinated (n = 8), TIV/Spn- (n=3), TIV/Spn+ (n=5), LAIV/Spn- (n=7) and LAIV/Spn+ (n=7) individuals. γδ, gamma-delta T cells; DN, double negative T cells. Nominal significance levels: ns = not significant; *p < 0.05; **p < 0.01; ***p < 0.001.

**Supp Figure 5.**
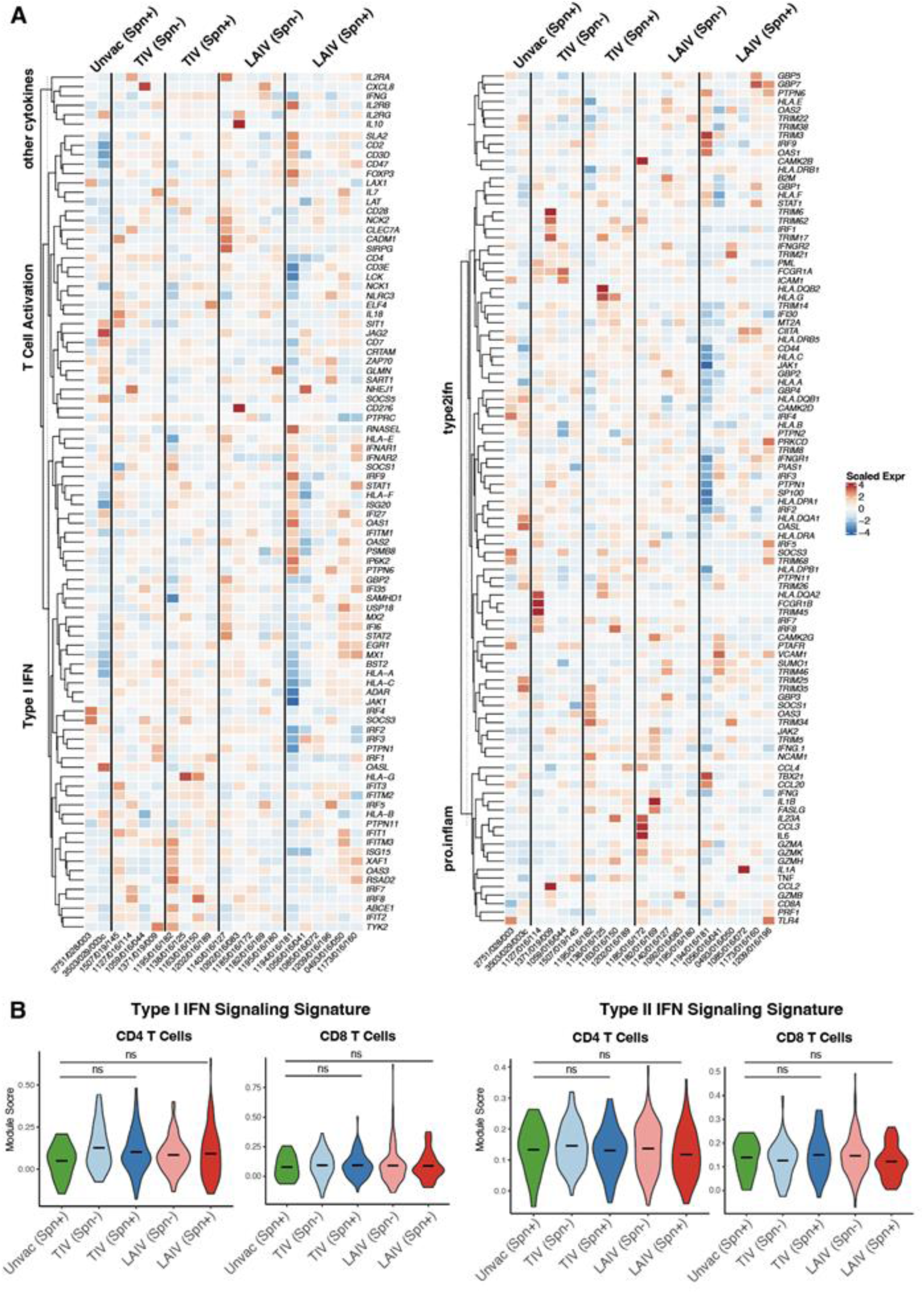
**A.** Heatmap of pro-inflammatory, T cell activation, and interferon-associated gene expressions across the 6 groups. **B.** Module scores of type I interferon and type II interferon signaling pathways in CD4+ (left) and CD8+ (right) T cells in single-cell level.

## References

1. Morens, D.M., Taubenberger, J.K., and Fauci, A.S. (2008). Predominant role of bacterial pneumonia as a cause of death in pandemic influenza: implications for pandemic influenza preparedness. J. Infect. Dis. 198, 962–970. 10.1086/591708.

2. Wright, A.K.A., Bangert, M., Gritzfeld, J.F., Ferreira, D.M., Jambo, K.C., Wright, A.D., Collins, A.M., and Gordon, S.B. (2013). Experimental Human Pneumococcal Carriage Augments IL-17A-dependent T-cell Defence of the Lung. PLOS Pathogens 9, e1003274. 10.1371/journal.ppat.1003274.

3. Ferreira, D.M., Neill, D.R., Bangert, M., Gritzfeld, J.F., Green, N., Wright, A.K.A., Pennington, S.H., Moreno, L.B., Moreno, A.T., Miyaji, E.N., et al. (2013). Controlled Human Infection and Rechallenge with Streptococcus pneumoniae Reveals the Protective Efficacy of Carriage in Healthy Adults. American Journal of Respiratory and Critical Care Medicine 187, 855. 10.1164/rccm.201212-2277OC.

4. Simell, B., Lahdenkari, M., Reunanen, A., Käyhty, H., and Väkeväinen, M. (2008). Effects of ageing and gender on naturally acquired antibodies to pneumococcal capsular polysaccharides and virulence-associated proteins. Clin Vaccine Immunol 15, 1391–1397. 10.1128/CVI.00110-08.

5. Wilson, R., Cohen, J.M., Reglinski, M., Jose, R.J., Chan, W.Y., Marshall, H., de Vogel, C., Gordon, S., Goldblatt, D., Petersen, F.C., et al. (2017). Naturally Acquired Human Immunity to Pneumococcus Is Dependent on Antibody to Protein Antigens. PLoS Pathog 13, e1006137. 10.1371/journal.ppat.1006137.

6. Thors, V., Christensen, H., Morales-Aza, B., Vipond, I., Muir, P., and Finn, A. (2016). The Effects of Live Attenuated Influenza Vaccine on Nasopharyngeal Bacteria in Healthy 2 to 4 Year Olds. A Randomized Controlled Trial. Am J Respir Crit Care Med 193, 1401–1409. 10.1164/rccm.201510-2000OC.

7. Jochems, S.P., de Ruiter, K., Solórzano, C., Voskamp, A., Mitsi, E., Nikolaou, E., Carniel, B.F., Pojar, S., German, E.L., Reiné, J., et al. (2019). Innate and adaptive nasal mucosal immune responses following experimental human pneumococcal colonization. J. Clin. Invest. 129, 4523–4538. 10.1172/JCI128865.

8. Hoft, D.F., Lottenbach, K.R., Blazevic, A., Turan, A., Blevins, T.P., Pacatte, T.P., Yu, Y., Mitchell, M.C., Hoft, S.G., and Belshe, R.B. (2017). Comparisons of the Humoral and Cellular Immune Responses Induced by Live Attenuated Influenza Vaccine and Inactivated Influenza Vaccine in Adults. Clin Vaccine Immunol 24. 10.1128/CVI.00414-16.

9. Dunne, E.M., Hua, Y., Salaudeen, R., Hossain, I., Ndiaye, M., Ortika, B.D., Mulholland, E.K., Hinds, J., Manna, S., Mackenzie, G.A., et al. (2022). Insights Into Pneumococcal Pneumonia Using Lung Aspirates and Nasopharyngeal Swabs Collected From Pneumonia Patients in The Gambia. J Infect Dis 225, 1447–1451. 10.1093/infdis/jiaa186.

10. Mitsi, E., Carniel, B., Reiné, J., Rylance, J., Zaidi, S., Soares-Schanoski, A., Connor, V., Collins, A.M., Schlitzer, A., Nikolaou, E., et al. (2020). Nasal Pneumococcal Density Is Associated with Microaspiration and Heightened Human Alveolar Macrophage Responsiveness to Bacterial Pathogens. Am J Respir Crit Care Med 201, 335–347. 10.1164/rccm.201903-0607OC.

11. Alveolar Macrophages (2018). Cellular Immunology 330, 86–90. 10.1016/j.cellimm.2018.01.005.

12. Rylance, J., de Steenhuijsen Piters, W.A.A., Mina, M.J., Bogaert, D., French, N., Ferreira, D.M., and EHPC-LAIV Study Group (2019). Two Randomized Trials of the Effect of Live Attenuated Influenza Vaccine on Pneumococcal Colonization. Am. J. Respir. Crit. Care Med. 199, 1160–1163. 10.1164/rccm.201811-2081LE.

13. Li, K., Neumann, K., Duhan, V., Namineni, S., Hansen, A.L., Wartewig, T., Kurgyis, Z., Holm, C.K., Heikenwalder, M., Lang, K.S., et al. (2019). The uric acid crystal receptor Clec12A potentiates type I interferon responses. Proc. Natl. Acad. Sci. U. S. A. 116, 18544–18549. 10.1073/pnas.1821351116.

14. Langfelder, P., and Horvath, S. (2008). WGCNA: an R package for weighted correlation network analysis. BMC Bioinformatics 9, 559. 10.1186/1471-2105-9-559.

15. Cheng, S., Li, Z., Gao, R., Xing, B., Gao, Y., Yang, Y., Qin, S., Zhang, L., Ouyang, H., Du, P., et al. (2021). A pan-cancer single-cell transcriptional atlas of tumor infiltrating myeloid cells. Cell 184, 792–809.e23. 10.1016/j.cell.2021.01.010.

16. Garcia-Alonso, L., Holland, C.H., Ibrahim, M.M., Turei, D., and Saez-Rodriguez, J. (2019). Benchmark and integration of resources for the estimation of human transcription factor activities. Genome Res. 29, 1363–1375. 10.1101/gr.240663.118.

17. O’Shea, J.J., Lahesmaa, R., Vahedi, G., Laurence, A., and Kanno, Y. (2011). Genomic views of STAT function in CD4+ T helper cell differentiation. Nat. Rev. Immunol. 11, 239–250. 10.1038/nri2958.

18. Yang, C., Mai, H., Peng, J., Zhou, B., Hou, J., and Jiang, D. (2020). STAT4: an immunoregulator contributing to diverse human diseases. Int. J. Biol. Sci. 16, 1575–1585. 10.7150/ijbs.41852.

19. Cambier, S., Gouwy, M., and Proost, P. (2023). The chemokines CXCL8 and CXCL12: molecular and functional properties, role in disease and efforts towards pharmacological intervention. Cellular & Molecular Immunology 20, 217–251. 10.1038/s41423-023-00974-6.

20. Zhou, C., Gao, Y., Ding, P., Wu, T., and Ji, G. (2023). The role of CXCL family members in different diseases. Cell Death Discov. 9, 212. 10.1038/s41420-023-01524-9.

21. Peters, J.M., Blainey, P.C., and Bryson, B.D. (2020). Consensus transcriptional states describe human mononuclear phagocyte diversity in the lung across health and disease. bioRxiv. 10.1101/2020.08.06.240424.

22. Liao, M., Liu, Y., Yuan, J., Wen, Y., Xu, G., Zhao, J., Cheng, L., Li, J., Wang, X., Wang, F., et al. (2020). Single-cell landscape of bronchoalveolar immune cells in patients with COVID-19. Nat. Med. 26, 842–844. 10.1038/s41591-020-0901-9.

23. Bischoff, P., Trinks, A., Wiederspahn, J., Obermayer, B., Pett, J.P., Jurmeister, P., Elsner, A., Dziodzio, T., Rückert, J.-C., Neudecker, J., et al. (2022). The single-cell transcriptional landscape of lung carcinoid tumors. Int. J. Cancer 150, 2058–2071. 10.1002/ijc.33995.

24. Chan, J.M., Quintanal-Villalonga, Á., Gao, V.R., Xie, Y., Allaj, V., Chaudhary, O., Masilionis, I., Egger, J., Chow, A., Walle, T., et al. (2021). Signatures of plasticity, metastasis, and immunosuppression in an atlas of human small cell lung cancer. Cancer Cell 39, 1479–1496.e18. 10.1016/j.ccell.2021.09.008.

25. Nilsson, A., Peters, J.M., Meimetis, N., Bryson, B., and Lauffenburger, D.A. (2022). Artificial neural networks enable genome-scale simulations of intracellular signaling. Nat. Commun. 13. 10.1038/s41467-022-30684-y.

26. Takeuchi, O., Hoshino, K., Kawai, T., Sanjo, H., Takada, H., Ogawa, T., Takeda, K., and Akira, S. (1999). Differential roles of TLR2 and TLR4 in recognition of gram-negative and gram-positive bacterial cell wall components. Immunity 11, 443–451. 10.1016/s1074-7613(00)80119-3.

27. Jing, J., Yang, I.V., Hui, L., Patel, J.A., Evans, C.M., Prikeris, R., Kobzik, L., O’Connor, B.P., and Schwartz, D.A. (2013). Role of macrophage receptor with collagenous structure in innate immune tolerance. J Immunol 190, 6360–6367. 10.4049/jimmunol.1202942.

28. Arredouani, M.S., Palecanda, A., Koziel, H., Huang, Y.-C., Imrich, A., Sulahian, T.H., Ning, Y.Y., Yang, Z., Pikkarainen, T., Sankala, M., et al. (2005). MARCO is the major binding receptor for unopsonized particles and bacteria on human alveolar macrophages. J Immunol 175, 6058–6064. 10.4049/jimmunol.175.9.6058.

29. Dai, X., Jayapal, M., Tay, H.K., Reghunathan, R., Lin, G., Too, C.T., Lim, Y.T., Chan, S.H., Kemeny, D.M., Floto, R.A., et al. (2009). Differential signal transduction, membrane trafficking, and immune effector functions mediated by FcgammaRI versus FcgammaRIIa. Blood 114, 318–327. 10.1182/blood-2008-10-184457.

30. Bowdish, D.M.E., Sakamoto, K., Kim, M.-J., Kroos, M., Mukhopadhyay, S., Leifer, C.A., Tryggvason, K., Gordon, S., and Russell, D.G. (2009). MARCO, TLR2, and CD14 are required for macrophage cytokine responses to mycobacterial trehalose dimycolate and Mycobacterium tuberculosis. PLoS Pathog 5, e1000474. 10.1371/journal.ppat.1000474.

31. Carniel, B.F., Marcon, F., Rylance, J., German, E.L., Zaidi, S., Reiné, J., Negera, E., Nikolaou, E., Pojar, S., Solórzano, C., et al. (2021). Pneumococcal colonization impairs mucosal immune responses to live attenuated influenza vaccine. JCI Insight 6. 10.1172/jci.insight.141088.

32. Mattorre, B., Caristi, S., Donato, S., Volpe, E., Faiella, M., Paiardini, A., Sorrentino, R., and Paladini, F. (2022). A short ERAP2 that binds IRAP is expressed in macrophages independently of gene variation. Int. J. Mol. Sci. 23, 4961. 10.3390/ijms23094961.

33. Raja, A., and Kuiper, J.J.W. (2023). Evolutionary immuno-genetics of endoplasmic reticulum aminopeptidase II (ERAP2). Genes Immun. 24, 295–302. 10.1038/s41435-023-00225-8.

34. Reddy, K.D., Rathnayake, S.N.H., Idrees, S., Boedijono, F., Xenaki, D., Padula, M.P., van den Berge, M., Faiz, A., and Oliver, B.G.G. (2025). A novel regulatory role for RPS4Y1 in inflammatory and fibrotic processes. Int. J. Mol. Sci. 26, 6213. 10.3390/ijms26136213.

35. Turner, P.J., Fleming, L., Saglani, S., Southern, J., Andrews, N.J., Miller, E., and SNIFFLE-4 Study Investigators (2020). Safety of live attenuated influenza vaccine (LAIV) in children with moderate to severe asthma. J. Allergy Clin. Immunol. 145, 1157–1164.e6. 10.1016/j.jaci.2019.12.010.

36. Duffy, J., Lewis, M., Harrington, T., Baxter, R., Belongia, E.A., Jackson, L.A., Jacobsen, S.J., Lee, G.M., Naleway, A.L., Nordin, J., et al. (2017). Live attenuated influenza vaccine use and safety in children and adults with asthma. Ann. Allergy Asthma Immunol. 118, 439–444. 10.1016/j.anai.2017.01.030.

37. Single-Cell Map of Diverse Immune Phenotypes in the Breast Tumor Microenvironment (2018). Cell 174, 1293–1308.e36. 10.1016/j.cell.2018.05.060.

38. Ashburner, M., Ball, C.A., Blake, J.A., Botstein, D., Butler, H., Cherry, J.M., Davis, A.P., Dolinski, K., Dwight, S.S., Eppig, J.T., et al. (2000). Gene ontology: tool for the unification of biology. The Gene Ontology Consortium. Nat Genet 25, 25–29. 10.1038/75556.

39. Gene Ontology Consortium, Aleksander, S.A., Balhoff, J., Carbon, S., Cherry, J.M., Drabkin, H.J., Ebert, D., Feuermann, M., Gaudet, P., Harris, N.L., et al. (2023). The Gene Ontology knowledgebase in 2023. Genetics 224. 10.1093/genetics/iyad031.

40. Salim, T., Sershen, C.L., and May, E.E. (2016). Investigating the Role of TNF-α and IFN-γ Activation on the Dynamics of iNOS Gene Expression in LPS Stimulated Macrophages. PLoS One 11. 10.1371/journal.pone.0153289.

41. Forman, H.J., and Torres, M. (2002). Reactive oxygen species and cell signaling: respiratory burst in macrophage signaling. Am. J. Respir. Crit. Care Med. 166, S4–S8. 10.1164/rccm.2206007.

42. Reeves, E., Colebatch-Bourn, A., Elliott, T., and Edwards, C.J. (2013). Function and genetics of ERAP1 and ERAP2: Implications for disease associations. *Human Immunology 74, 867–872.

43. Aegerter, H., Kulikauskaite, J., Crotta, S., Patel, H., Kelly, G., Hessel, E.M., Mack, M., Beinke, S., and Wack, A. (2020). Influenza-induced monocyte-derived alveolar macrophages confer prolonged antibacterial protection. Nat. Immunol. 21, 145–157. 10.1038/s41590-019-0568-x.

44. Mitsi, E., McLenaghan, D., Wolf, A.-S., Jones, S., Collins, A.M., Hyder-Wright, A.D., Goldblatt, D., Heyderman, R.S., Gordon, S.B., and Ferreira, D.M. (2022). Thirteen-Valent pneumococcal conjugate vaccine-induced immunoglobulin G (IgG) responses in serum associated with serotype-specific IgG in the lung. J. Infect. Dis. 225, 1626–1631. 10.1093/infdis/jiab331.

45. Lee, A., Floyd, K., Wu, S., Fang, Z., Tan, T.K., Froggatt, H.M., Powers, J.M., Leist, S.R., Gully, K.L., Hubbard, M.L., et al. (2024). BCG vaccination stimulates integrated organ immunity by feedback of the adaptive immune response to imprint prolonged innate antiviral resistance. Nat Immunol 25, 41–53. 10.1038/s41590-023-01700-0.

46. Zhang, H., Floyd, K., Fang, Z., Hoffmann, F.A., Lee, A., Froggatt, H.M., Bharj, G., Xie, X., Eppler, H.B., Santagata, J.M., et al. (2026). Mucosal vaccination in mice provides protection from diverse respiratory threats. Science 392, eaea1260. 10.1126/science.aea1260.

47. Bailey, J.I., Puritz, C.H., Senkow, K.J., Markov, N.S., Diaz, E., Jonasson, E., Yu, Z., Swaminathan, S., Lu, Z., Fenske, S., et al. (2024). Profibrotic monocyte-derived alveolar macrophages are expanded in patients with persistent respiratory symptoms and radiographic abnormalities after COVID-19. Nat Immunol 25, 2097–2109. 10.1038/s41590-024-01975-x.

48. Madissoon, E., Oliver, A.J., Kleshchevnikov, V., Wilbrey-Clark, A., Polanski, K., Richoz, N., Ribeiro Orsi, A., Mamanova, L., Bolt, L., Elmentaite, R., et al. (2023). A spatially resolved atlas of the human lung characterizes a gland-associated immune niche. Nat Genet 55, 66–77. 10.1038/s41588-022-01243-4.

49. Xu, C., Prete, M., Webb, S., Jardine, L., Stewart, B.J., Hoo, R., He, P., Meyer, K.B., and Teichmann, S.A. (2023). Automatic cell-type harmonization and integration across Human Cell Atlas datasets. Cell 186, 5876–5891.e20. 10.1016/j.cell.2023.11.026.

50. Domínguez Conde, C., Xu, C., Jarvis, L.B., Rainbow, D.B., Wells, S.B., Gomes, T., Howlett, S.K., Suchanek, O., Polanski, K., King, H.W., et al. (2022). Cross-tissue immune cell analysis reveals tissue-specific features in humans. Science 376, eabl5197. 10.1126/science.abl5197.

51. Stuart, T., Butler, A., Hoffman, P., Hafemeister, C., Papalexi, E., Mauck, W.M., 3rd, Hao, Y., Stoeckius, M., Smibert, P., and Satija, R. (2019). Comprehensive Integration of Single-Cell Data. Cell 177, 1888–1902.e21. 10.1016/j.cell.2019.05.031.

52. Hao, Y., Stuart, T., Kowalski, M.H., Choudhary, S., Hoffman, P., Hartman, A., Srivastava, A., Molla, G., Madad, S., Fernandez-Granda, C., et al. (2024). Dictionary learning for integrative, multimodal and scalable single-cell analysis. Nat Biotechnol 42, 293–304. 10.1038/s41587-023-01767-y.

53. GitHub - maiermarco/DirichletReg GitHub. https://github.com/maiermarco/DirichletReg.

54. Finak, G., McDavid, A., Yajima, M., Deng, J., Gersuk, V., Shalek, A.K., Slichter, C.K., Miller, H.W., McElrath, M.J., Prlic, M., et al. (2015). MAST: a flexible statistical framework for assessing transcriptional changes and characterizing heterogeneity in single-cell RNA sequencing data. Genome Biol. 16, 278. 10.1186/s13059-015-0844-5.

55. Holland, C.H., Tanevski, J., Perales-Patón, J., Gleixner, J., Kumar, M.P., Mereu, E., Joughin, B.A., Stegle, O., Lauffenburger, D.A., Heyn, H., et al. (2020). Robustness and applicability of transcription factor and pathway analysis tools on single-cell RNA-seq data. Genome Biol. 21, 36. 10.1186/s13059-020-1949-z.

56. Dimitrov, D., Türei, D., Garrido-Rodriguez, M., Burmedi, P.L., Nagai, J.S., Boys, C., Ramirez Flores, R.O., Kim, H., Szalai, B., Costa, I.G., et al. (2022). Comparison of methods and resources for cell-cell communication inference from single-cell RNA-Seq data. Nat Commun 13, 3224. 10.1038/s41467-022-30755-0.

57. Browaeys, R., Saelens, W., and Saeys, Y. (2020). NicheNet: modeling intercellular communication by linking ligands to target genes. Nat Methods 17, 159–162. 10.1038/s41592-019-0667-5.

58. Jin, S., Plikus, M.V., and Nie, Q. (2025). CellChat for systematic analysis of cell-cell communication from single-cell transcriptomics. Nat Protoc 20, 180–219. 10.1038/s41596-024-01045-4.

